# Evaluation of a SARS-CoV-2 Vaccine NVX-CoV2373 in Younger and Older Adults

**DOI:** 10.1101/2021.02.26.21252482

**Authors:** Neil Formica, Raburn Mallory, Gary Albert, Michelle Robinson, Joyce S. Plested, Iksung Cho, Andreana Robertson, Filip Dubovsky, Gregory M. Glenn, for the 2019nCoV-101 Study Group

## Abstract

**Background:** NVX-CoV2373 is a recombinant severe acute respiratory coronavirus 2 (rSARS-CoV-2) nanoparticle vaccine composed of trimeric full-length SARS-CoV-2 spike glycoproteins and Matrix-M1 adjuvant.

**Methods:** The phase 2 component of our randomized, placebo-controlled, phase 1-2 trial was designed to identify which dosing regimen of NVX-CoV2373 should move forward into late phase studies in younger (18-59 years) and older (60-84 years) participants and was based on immunogenicity and safety data through day 35 (14 days after the second dose). Participants were randomly assigned to receive either one or two intramuscular doses of 5-µg or 25-µg NVX-CoV2373 or placebo, 21 days apart. Primary endpoints were immunoglobulin G (IgG) anti-spike protein response, 7-day solicited reactogenicity, and unsolicited adverse events. A key secondary endpoint was wild-type virus neutralizing antibody response.

**Results:** After randomization, approximately 250 participants each were assigned to one of four vaccine groups or placebo. Of these, approximately 45% were older participants. Reactogenicity was predominantly mild to moderate in severity and of short duration (median <3 days) after first and second vaccination with NVX-CoV2373, with higher frequencies and intensity after second vaccination and with the higher dose, and occurred less frequently and was of lower intensity in older participants. The two-dose regimen of 5-µg NVX-CoV2373 induced robust geometric mean titer (GMT) IgG anti-spike protein (65,019 and 28,137 EU/mL) and wild-type virus neutralizing antibody (2201 and 981 titers) responses in younger and older participants, respectively, with seroconversion rates of 100% in both age groups. Neutralizing antibody responses exceeded those seen in convalescent sera for both age groups.

**Conclusions:** The study confirmed that the two-dose regimen of 5-µg NVX-CoV2373 is highly immunogenic and well tolerated in both younger and older participants.

(Funded by the Coalition for Epidemic Preparedness Innovations; ClinicalTrials.gov number: NCT04368988).

## INTRODUCTION

The coronavirus disease 2019 (COVID-19) pandemic caused by the severe acute respiratory syndrome coronavirus 2 (SARS-CoV-2) has continued to spread rapidly throughout the world, with over 109 million confirmed cases and over 2.4 million deaths globally as of February 18, 2021.^1^ Mutated SARS-CoV-2 variants with enhanced transmissibility emerged and established themselves as clinically dominant in the United Kingdom and South Africa in late 2020 and risk spreading globally. Although several vaccine products against SARS-CoV-2 are authorized for use in some countries, their supplies are limited and their effectiveness against mutated variants has yet to be demonstrated. Therefore, there is a growing health and economic need for safe and efficacious vaccines to help prevent the spread of COVID-19 (including its variants) throughout the world.

NVX-CoV2373 contains Matrix-M1 adjuvant^2^ and a recombinant SARS-CoV-2 (rSARS-CoV-2) nanoparticle vaccine,^3^ constructed from the full-length (i.e., including the transmembrane domain), wild-type SARS-CoV-2 spike glycoprotein, which mediates viral attachment to the human angiotensin-converting enzyme 2 (hACE2) receptor of host cells and serves as a target for development of antibodies and vaccines.^4,5^ Targeting the SARS-CoV-2 spike protein has recently been shown to be highly effective in preventing COVID-19 in clinical trials, which has formed the basis for emergency use of several vaccine candidates.^6,7^ This is encouraging news for the development of NVX-CoV2373.

The phase 1 component of the phase 1-2 trial showed that two-dose regimens of 5-µg and 25-µg rSARS-CoV-2 with 50-µg Matrix-M1 adjuvant (mixed prior to use) in participants 18 to 59 years were well tolerated and immunogenic by both immunoglobulin G (IgG) anti-spike protein enzyme-linked immunosorbent assay (ELISA) and fit for purpose wild-type viral microneutralization (MN) assay with an inhibitory concentration of >99% (MN_>99%_), with responses that exceeded levels seen in a panel of COVID-19 convalescent serum samples.^3^ Importantly, the Matrix-M1 adjuvant was dose sparing and also induced strong/high levels of CD4+ effector memory T-cell responses that were also biased toward a Th1 phenotype that may play a role in reducing the theoretical possibility of antibody-dependent enhancement (ADE) of SARS-CoV-2 infection.^8^

Here, we report on the safety and immunogenicity using qualified and validated immunoassays from the phase 2 component of the randomized, placebo-controlled, observer-blinded, phase 1-2 trial to confirm the optimal dose regimen of NVX-CoV2373 in younger and older adult participants.

## METHODS

### Trial Design and Oversight

Our phase 2 trial was conducted at nine sites in Australia and eight sites in the United States. Eligible participants were men and non-pregnant women 18 to 84 years of age with a body mass index (the weight in kilograms divided by the square of the height in meters) of 17 to 35. Participants with underlying medical conditions could be enrolled if the conditions were judged clinically to be stable, as were participants with confirmed COVID-19 presenting with mild symptomatology. Details of the trial design, conduct, oversight, and analyses are provided in the protocol and statistical analysis plan (to be made available at publication). All participants provided written informed consent prior to trial enrollment.

Approximately 750 participants were planned to be randomized in each country. Eligible participants were randomly assigned (1:1:1:1:1) in a blinded manner to one of five vaccine groups (groups A, B, C, D, and E) according to pre-generated randomization schedules with two-factor, two-level stratification employed (ages 18-59 and 60-84 years; study site) (Fig. 1). As a safety measure, enrollment of older participants was initially limited to approximately 50 participants in each of the five vaccine groups until reactogenicity was observed for 5 days with no safety concerns.

**Figure 1.**
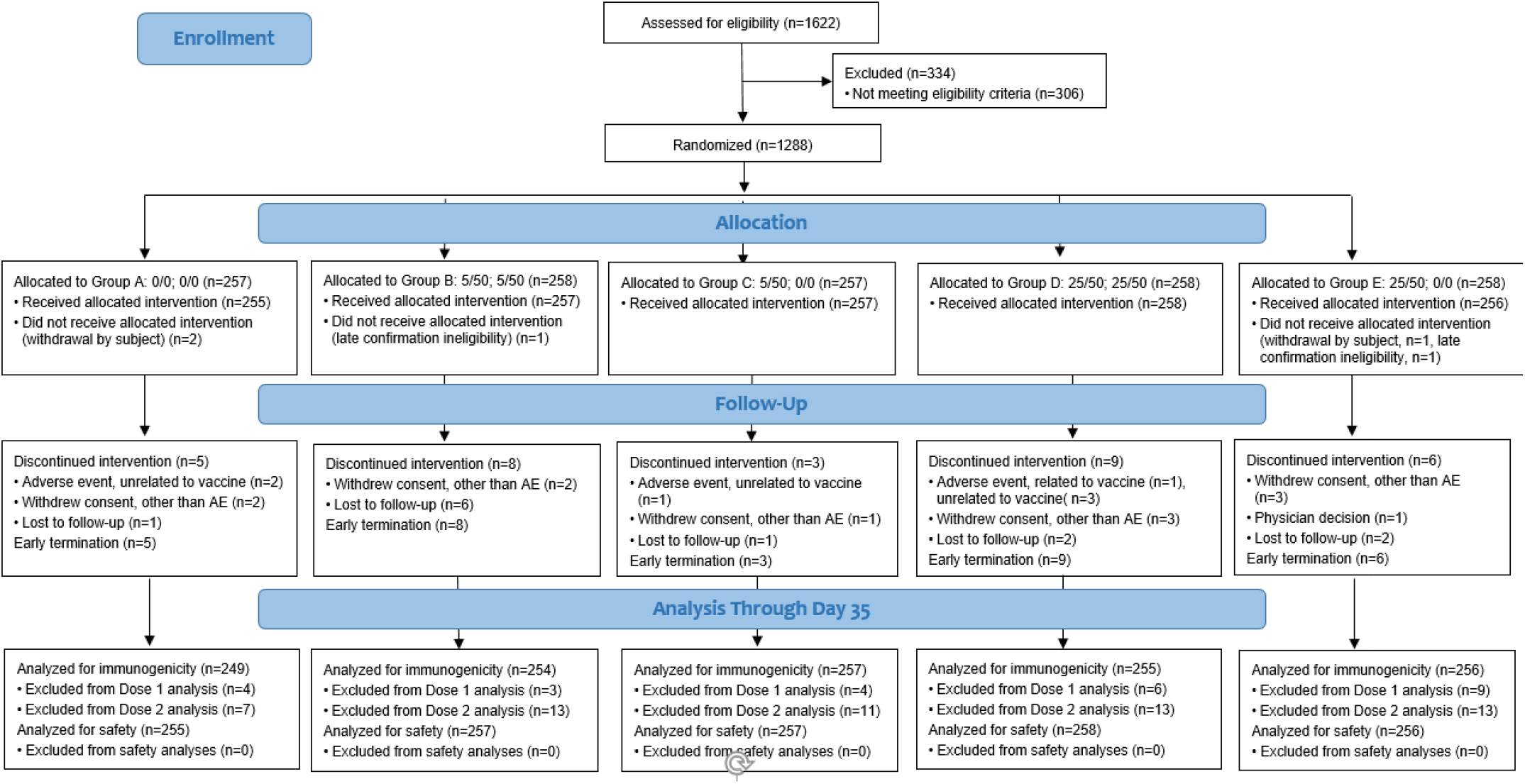
CONSORT Flow Diagram.

Each participant received two intramuscular injections of 5-µg (low dose) or 25-µg (high dose) NVX-CoV2373 and/or placebo in a one-dose (5-µg or 25-µg followed by placebo) or two-dose (5-µg followed by 5-µg or 25-µg followed by 25-µg) regimen, 21 days apart. Participants initially randomized to treatment groups B and C will be re-randomized at day 189 in a 1:1 ratio to receive either a dose of vaccine (B2 and C2) or placebo (B1 or C1); these results will be reported once available. Participants and trial site personnel managing the conduct of the trial remained blinded to vaccine assignment. Vaccination pause rules were in place to monitor participants’ safety (see Supplementary Appendix).

The trial was designed by Novavax, Inc. (Gaithersburg, Maryland, USA), with funding support from the Coalition for Epidemic Preparedness Innovations. The trial protocol was approved by the Alfred Hospital Human Research Ethics Committee (Melbourne, Australia) and Advarra Central Institutional Review Board (Colombia, Maryland, USA) and was performed in accordance with the International Conference on Harmonisation, Good Clinical Practice guidelines. Safety oversight for the older adult lead-in enrollment cohort and specific vaccination pause rules were supported by an independent safety monitoring committee. The authors assume responsibility for the accuracy and completeness of the data and analyses, as well as for the fidelity of the trial.

### Trial Procedures: Safety Assessments

Reactogenicity was assessed within 7 days after each dose of trial vaccine, as prompted and recorded by an electronic participant diary. Predefined solicited local (injection site) reactogenicity included pain, tenderness, erythema, and swelling; systemic reactogenicity included fever, nausea or vomiting, headache, fatigue, malaise, myalgia, and arthralgia. Reactogenicity severity was graded according to the Food and Drug Administration (FDA) toxicity grading scale with minor modifications (Table S1).^9^ Unsolicited adverse events were assessed at each study visit and coded by preferred term and system organ class according to the *Medical Dictionary for Regulatory Activities* (MedDRA), version 23.0, and summarized by treatment group, clinical severity (mild, moderate, and severe) and causality (not related and related). These events included adverse events of special interest relevant to COVID-19 (Table S2)^10,11^ and to potential immune-mediated medical conditions (Table S3).

### Immunogenicity Assessments

A validated ELISA assay (Novavax, Inc., Gaithersburg, Maryland, USA) was used to measure IgG anti-spike protein levels specific for rSARS-CoV-2 protein antigens at days 0 (baseline), 21, and 35, with seropositivity defined as a titer above the lower limit of quantification (200 ELISA units per milliliter). 360biolabs (Melbourne, Australia) used a qualified MN assay with an inhibitory concentration of 50% (MN_50%_) to measure neutralizing antibodies specific to wild-type virus SARS-CoV-2 at the same time points, with seropositivity defined as titer above the lower limit of quantification (titer of 20). An ad hoc analysis was conducted, comparing the neutralizing antibody response to a sample panel of convalescent sera in patients with a median age of 55.^3^

### Statistical Analysis

The decision on which dosing regimen of NVX-CoV2373 to move forward into late-phase studies in both younger and older adult participants was based on the totality of the immunogenicity and safety data rather than any individual measurement. No multiplicity adjustment was made where multiple vaccine groups and endpoints were being evaluated. With at least 150 participants in each vaccine group, there was more than adequate power to detect differences in IgG anti-spike protein responses and safety data between vaccine groups. Most endpoints were summarized using descriptive statistics, with 95% confidence intervals (CIs) added based on the t-distribution of the log-transformed values.

## RESULTS

### Trial Population

Between August 24, 2020, and September 25, 2020, a total of 1288 participants underwent randomization at 17 sites in Australia and the United States in the phase 2 component of the trial (Fig. S1). A total of 1283 participants received the initial injection, with 255 in the placebo group (group A), 257 in the 5-µg two-dose regimen (group B), 257 in the 5-µg one-dose regimen (group C), 258 in the 25-µg two-dose regimen (group D), and 256 in the 25-µg two-dose regimen (group E). A total of 1256 participants received both injections (250, 254, 255, 251, and 246 in groups A, B, C, D, and E, respectively). Demographic characteristics of the participants are presented in Table 1. The mean age was 52 years, with 45% in the older age group. Among the 1283 participants, 49% were male, 87% were White, 4% were of Hispanic or Latino origin, and 2% had a positive baseline SARS-CoV-2 serostatus.

**Table 1.**
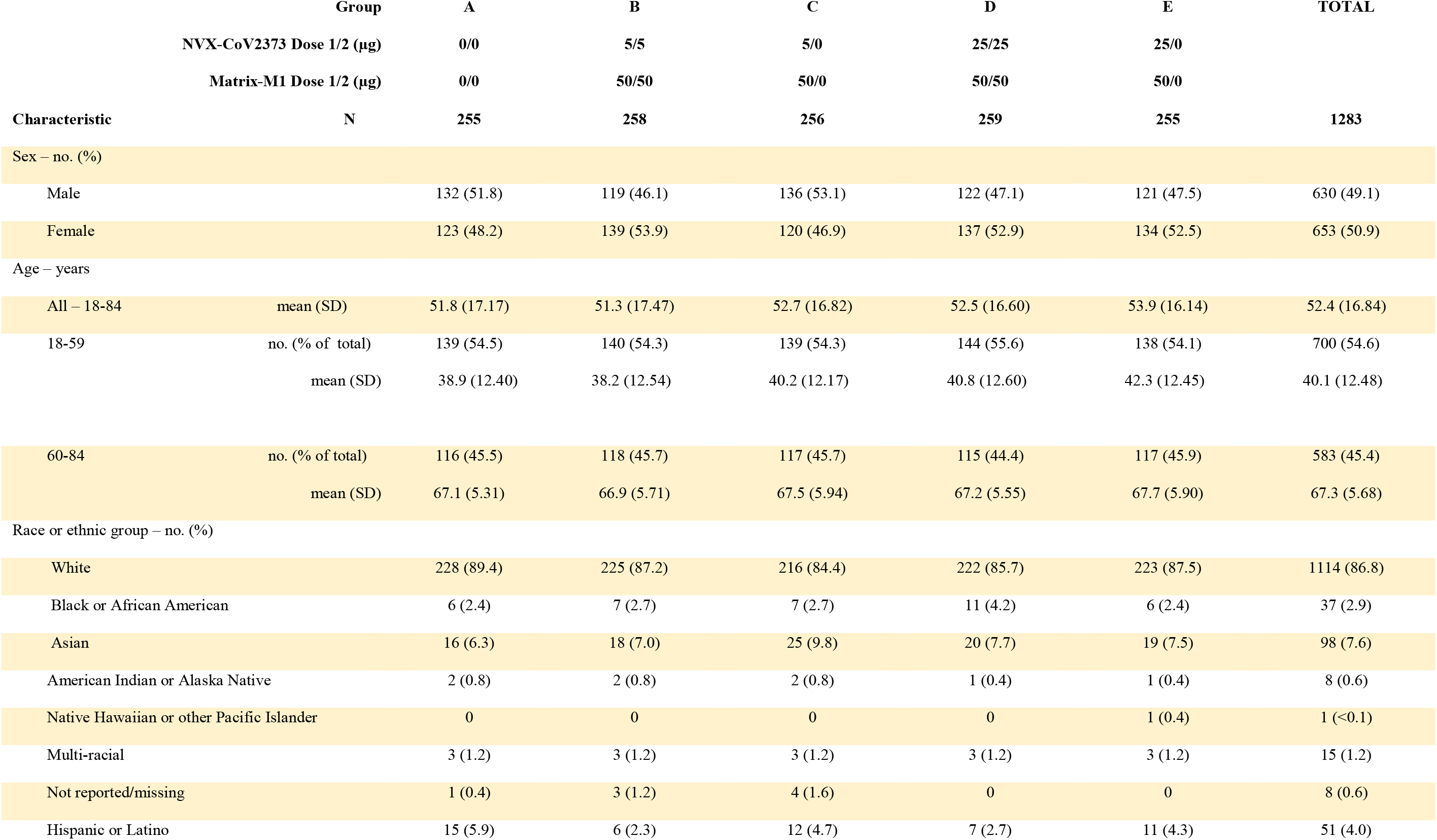
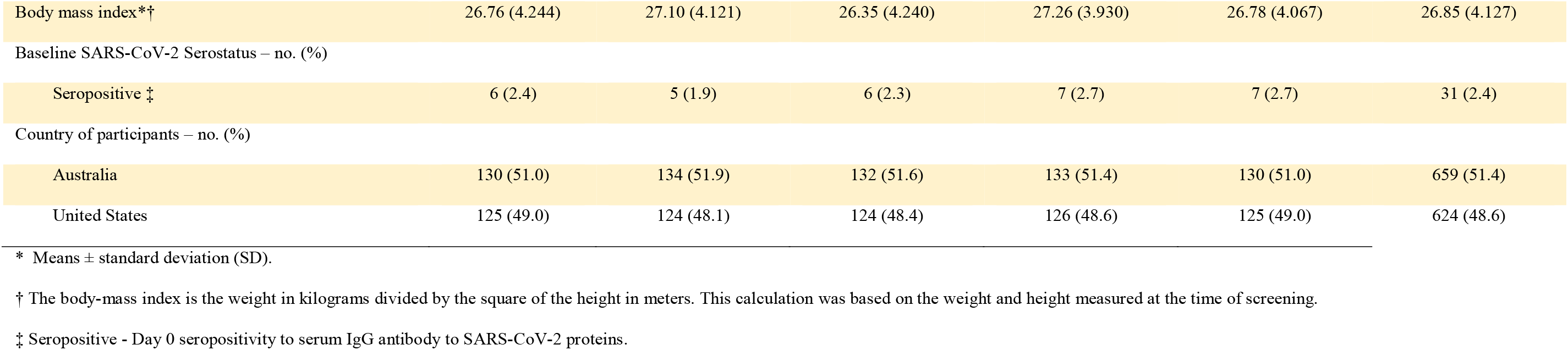
Demographic Characteristics of the Participants in the NVX-CoV2373 Phase 2 Trial 2019nCoV-101 at Enrollment.

|  | Group | A | B | C | D | E | TOTAL |
| --- | --- | --- | --- | --- | --- | --- | --- |
|  | NVX-CoV2373 Dose 1/2 (µg) | 0/0 | 5/5 | 5/0 | 25/25 | 25/0 |  |
|  | Matrix-M1 Dose 1/2 (µg) | 0/0 | 50/50 | 50/0 | 50/50 | 50/0 |  |
| Characteristic | N | 255 | 258 | 256 | 259 | 255 | 1283 |
| Sex – no. (%) |  |  |  |  |  |  |  |
| Male |  | 132 (51.8) | 119 (46.1) | 136 (53.1) | 122 (47.1) | 121 (47.5) | 630 (49.1) |
| Female |  | 123 (48.2) | 139 (53.9) | 120 (46.9) | 137 (52.9) | 134 (52.5) | 653 (50.9) |
| Age – years |  |  |  |  |  |  |  |
| All – 18-84 | mean (SD) | 51.8 (17.17) | 51.3 (17.47) | 52.7 (16.82) | 52.5 (16.60) | 53.9 (16.14) | 52.4 (16.84) |
| 18-59 | no. (% of total) | 139 (54.5) | 140 (54.3) | 139 (54.3) | 144 (55.6) | 138 (54.1) | 700 (54.6) |
|  | mean (SD) | 38.9 (12.40) | 38.2 (12.54) | 40.2 (12.17) | 40.8 (12.60) | 42.3 (12.45) | 40.1 (12.48) |
| 60-84 | no. (% of total) | 116 (45.5) | 118 (45.7) | 117 (45.7) | 115 (44.4) | 117 (45.9) | 583 (45.4) |
|  | mean (SD) | 67.1 (5.31) | 66.9 (5.71) | 67.5 (5.94) | 67.2 (5.55) | 67.7 (5.90) | 67.3 (5.68) |
| Race or ethnic group – no. (%) |  |  |  |  |  |  |  |
| White |  | 228 (89.4) | 225 (87.2) | 216 (84.4) | 222 (85.7) | 223 (87.5) | 1114 (86.8) |
| Black or African American |  | 6 (2.4) | 7 (2.7) | 7 (2.7) | 11 (4.2) | 6 (2.4) | 37 (2.9) |
| Asian |  | 16 (6.3) | 18 (7.0) | 25 (9.8) | 20 (7.7) | 19 (7.5) | 98 (7.6) |
| American Indian or Alaska Native |  | 2 (0.8) | 2 (0.8) | 2 (0.8) | 1 (0.4) | 1 (0.4) | 8 (0.6) |
| Native Hawaiian or other Pacific Islander |  | 0 | 0 | 0 | 0 | 1 (0.4) | 1 (<0.1) |
| Multi-racial |  | 3 (1.2) | 3 (1.2) | 3 (1.2) | 3 (1.2) | 3 (1.2) | 15 (1.2) |
| Not reported/missing |  | 1 (0.4) | 3 (1.2) | 4 (1.6) | 0 | 0 | 8 (0.6) |
| Hispanic or Latino |  | 15 (5.9) | 6 (2.3) | 12 (4.7) | 7 (2.7) | 11 (4.3) | 51 (4.0) |
|  | NVX-CoV2373 Dose 1/2 (µg) | 0/0 | 5/5 | 5/0 | 25/25 | 25/0 |  |
|  | Matrix-M1 Dose 1/2 (µg) | 0/0 | 50/50 | 50/0 | 50/50 | 50/0 |  |
| Characteristic | N | 255 | 258 | 256 | 259 | 255 | 1283 |
| Body mass index*† |  | 26.76 (4.244) | 27.10 (4.121) | 26.35 (4.240) | 27.26 (3.930) | 26.78 (4.067) | 26.85 (4.127) |
| Baseline SARS-CoV-2 Serostatus – no. (%) |  |  |  |  |  |  |  |
| Seropositive ‡ |  | 6 (2.4) | 5 (1.9) | 6 (2.3) | 7 (2.7) | 7 (2.7) | 31 (2.4) |
| Country of participants – no. (%) |  |  |  |  |  |  |  |
| Australia |  | 130 (51.0) | 134 (51.9) | 132 (51.6) | 133 (51.4) | 130 (51.0) | 659 (51.4) |
| United States |  | 125 (49.0) | 124 (48.1) | 124 (48.4) | 126 (48.6) | 125 (49.0) | 624 (48.6) |

\* Means ± standard deviation (SD).
† The body-mass index is the weight in kilograms divided by the square of the height in meters. This calculation was based on the weight and height measured at the time of screening.
‡ Seropositive - Day 0 seropositivity to serum IgG antibody to SARS-CoV-2 proteins.

### Safety Outcomes

#### Solicited Local Reactogenicity

Across both age groups, solicited local adverse events were more predominant in the NVX-CoV2373 groups than in the placebo group following each vaccination (Fig. 2 and Table S7). Following first vaccination of 5-µg (groups B and C combined) and 25-µg (groups D and E combined) NVX-CoV2373, the most frequent solicited local adverse events across both age groups were tenderness (48% and 59%, respectively) and pain (27% and 38%). The frequencies of these events were higher among younger participants (61% and 68% for tenderness; 36% and 48% for pain) and lower among older participants (33% and 49% for tenderness; 17% and 25% for pain). Tenderness and pain were predominantly grade 1-2 in younger participants and grade 1 in older participants and of short duration (median of 1 day for pain and 2 days for tenderness) across both age groups. Grade 3 solicited local adverse events were reported in one low-dose (<1%) and two high-dose recipients (1%) among younger participants and in one high-dose recipient (<1%) among older participants; there were no grade 4 events.

**Figure 2.**
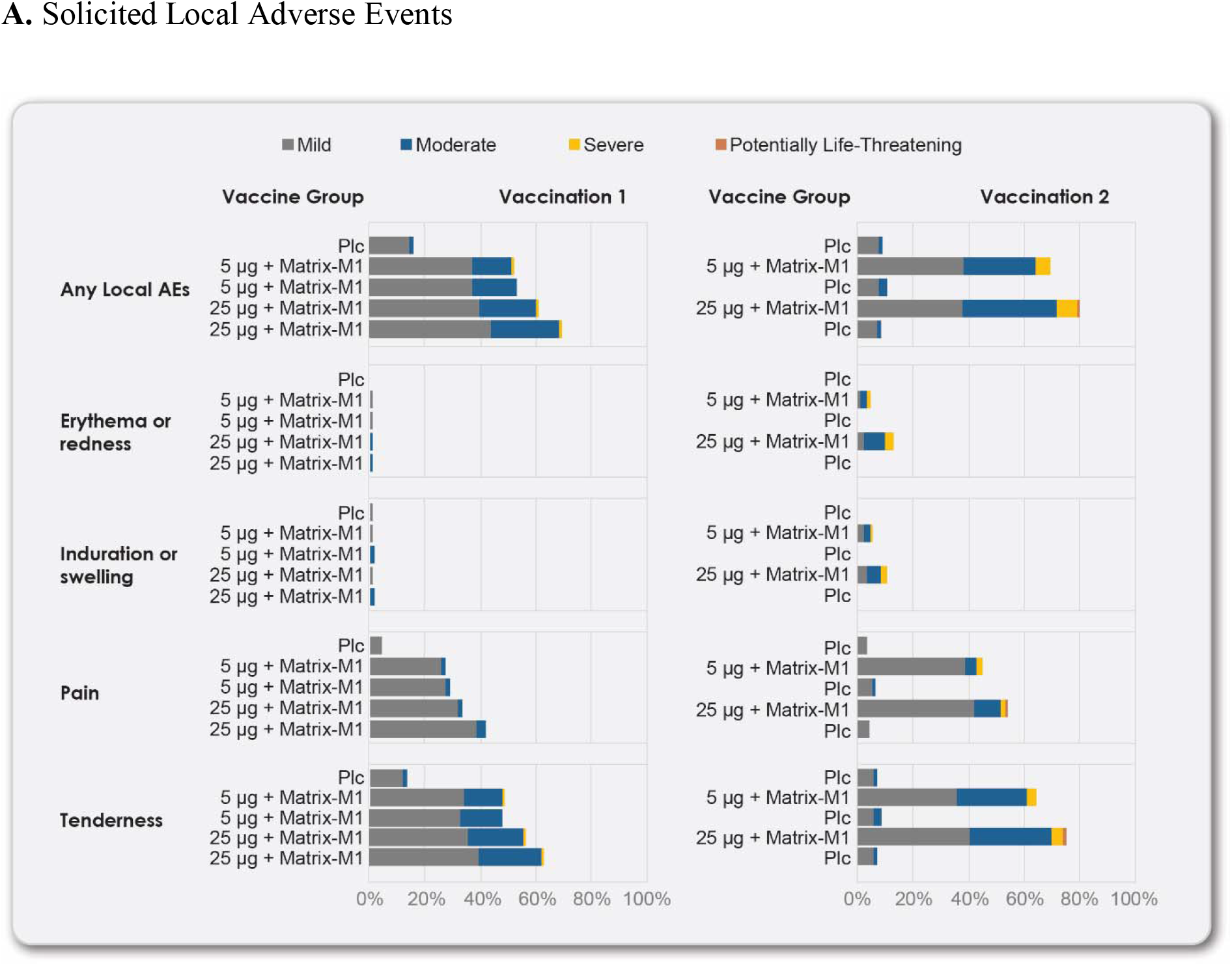
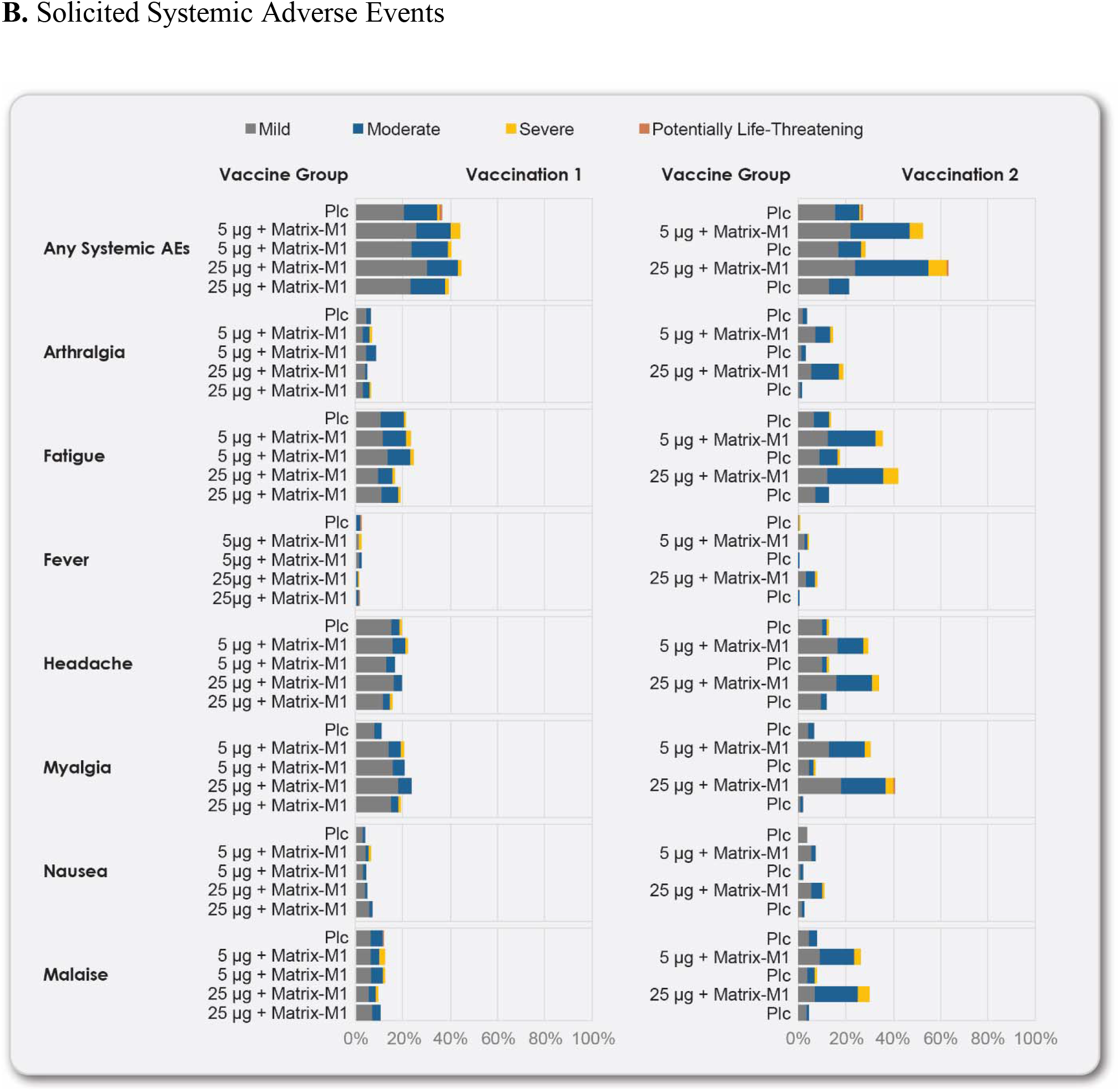
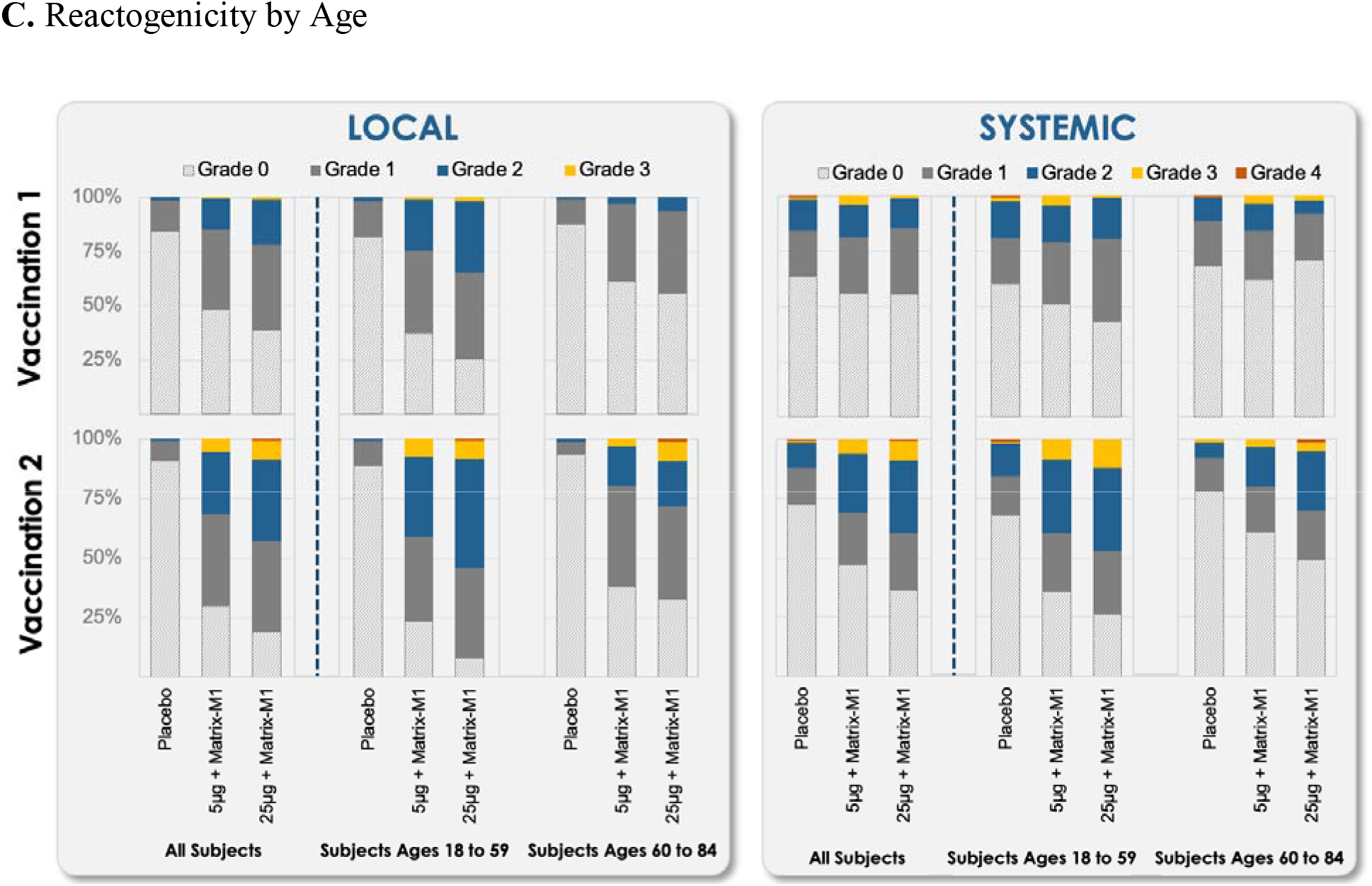
Solicited Local and Systemic Adverse Events. The percentage of participants in each vaccine group (groups A, B, C, D, and E) with adverse events according to the maximum toxicity grade (mild, moderate, severe, or potentially life-threatening) during the 7 days after each vaccination is plotted for solicited local (Panel A) and systemic (Panel B) adverse events and for both local and systemic events by age group (Panel C). Grade 4 (potentially life-threatening) events met this criteria by emergency department attendances. (See Table S4 for complete safety data on all participants.)

Following second vaccination of 5-µg (group B) and 25-µg (group D) NVX-CoV2373, the most frequent solicited local adverse events across both age groups were tenderness (65% and 76%) and pain (46% and 55%). The frequencies of these events were higher among younger participants (74% and 88% for tenderness; 50% and 67% for pain) and lower among older participants (55% and 62% for tenderness and 41% and 39% for pain). These events were predominantly grade 1 or 2 and of short duration (median of 2 days) across both age groups. For vaccine participants who received placebo as the second injection, frequencies of solicited local adverse events were similar to placebo. Grade 3 solicited local adverse events were reported in 10 low-dose recipients (7%) and 10 high-dose recipients (7%) among younger participants and in three low-dose recipients (3%) and nine high-dose recipients (8%) among older participants; grade 4 events were reported in one high-dose recipient (1%) in each age group.

#### Solicited Systemic Reactogenicity

Across both age groups, solicited systemic adverse events other than muscle pain were reported at similar frequencies between the NVX-CoV2373 and placebo groups following first vaccination; however, solicited systemic adverse events were more predominant in the NVX-CoV2373 groups than in the placebo group following second vaccination (Fig. 2 and Table S7). Following first vaccination of 5-µg (groups B and C combined) and 25-µg (groups D and E combined) NVX-CoV2373, muscle pain (20% and 21%) was predominantly grade 1 and of short duration (median of 1 day) across both age groups. The frequency of this event was higher among younger participants (25% and 27%) and lower among older participants (15% and 14%). Fever was reported in less than 2% of vaccine recipients across both age groups, with grade 3 fever reported in three low-dose recipients (1%) and one high-dose recipient (<1%) and a grade 4 fever reported in one high-dose (<1%) and one placebo (<1%) recipient. Grade 3 solicited systemic adverse events were reported in two placebo recipients (1%), seven low-dose recipients (3%), and four high-dose recipients (1%) among younger participants, with a grade 4 event reported in one placebo recipient (1%). Among older participants, grade 3 solicited systemic adverse events occurred in six low-dose recipients (3%) and two high-dose recipients (1%) and grade 4 events were reported in one placebo recipient (1%) and one high-dose recipient (1%).

Following second vaccination of 5-µg (group B) and 25-µg (group D) NVX-CoV2373, the most frequent solicited systemic adverse events were fatigue (36% and 43%), muscle pain (31% and 41%), headache (30% and 34%), and malaise (26% and 30%). The frequencies of these events were higher among younger participants (46% and 50% for fatigue, 40% and 50% for muscle pain, 42% and 41% for headache, and 31% and 41% for malaise) and lower among older participants (23% and 33% for fatigue, 20% and 29% for muscle pain, 15% and 26% for headache, and 20% and 16% for malaise). These events were predominantly grade 1 or 2 and of short duration (median of 1 day) across both age groups. For vaccine participants who received placebo as the second injection, frequencies of solicited systemic adverse events were similar to placebo. Fever was reported in one placebo recipient (1%), nine low-dose recipients (7%), and 16 high-dose recipients (12%) among younger participants, with grade 3 fever reported in one low-dose recipient (1%) and two high-dose recipients and grade 4 fever reported in one placebo recipient (1%). Among older participants, fever was reported in one placebo recipient (1%), two low-dose recipients (2%), and four high-dose recipients (4%), with no grade 3 or 4 fever. Grade 3 solicited systemic adverse events were reported in one placebo recipient (1%), 11 low-dose recipients (8%), and 16 high-dose recipients (12%) among younger participants and a grade 4 event was reported in one placebo recipient (<1%). Among older participants, grade 3 solicited systemic adverse events were reported in one placebo recipient (1%), three low-dose recipients (3%), and four high-dose recipients (4%) and a grade 4 event was reported in one high-dose recipient (1%).

### Unsolicited Adverse Events

Unsolicited adverse events were reported in 42 placebo recipients (17%), 86 low-dose recipients (17%), and 95 high-dose recipients (18%), with similar distributions across both age groups (Table S5). Unsolicited adverse events were predominantly mild across both age groups, with severe events reported in three placebo recipients (1%), five low-dose recipients (1%), and one high-dose recipient (<1%). One low-dose recipient (<1%) had a related severe unsolicited adverse event (acute colitis) that also met the criteria for a serious adverse event.

Seven participants discontinued the trial due to an unsolicited adverse event, including two placebo recipients (non-Hodgkin lymphoma; atrial fibrillation); one low-dose recipient (urinary incontinence); and four high-dose recipients (arthralgia; pyrexia, myalgia, and malaise (related); dermatitis; and atrial fibrillation). Nine serious adverse events were reported: one case of acute colitis, assessed as related by the investigator; one case of atrial fibrillation, assessed as not related by the investigator due to underlying cardiac disease; one case of vertigo (not related); one case of wrist fracture (not related); one case of non-Hodgkin’s lymphoma (not related); one case of animal bite (not related); one case of acute myocardial infarction, assessed as not related due to underlying risk factors (hypertension, type 2 diabetes, and hypercholesterolemia); one case of multiple sclerosis that was assessed as related by the investigator (placebo); and one case of lumbar spinal stenosis (not related). One placebo recipient had an adverse event of special interest associated with a potential immune-mediated medical condition (multiple sclerosis), and there were no adverse events of special interest associated with COVID-19. Details regarding all safety data are provided in the Supplementary Appendix.

### Immunogenicity Outcomes

#### Anti-Spike Protein Binding IgG Response

Anti-spike protein binding IgG geometric mean titers (GMTs) (reported as EU/mL) at day 35 were 44,421 [95% CI, 37,929 to 52,024) and 46,459 (95% CI, 40,839 to 52,853) for the two-dose regimens of 5-µg and 25-µg NVX-CoV2373, respectively, in participants 18 to 84 years of age who received full vaccination regardless of baseline serostatus (Table S6). This equated to geometric mean fold rises (GMFRs) of 386 and 385 relative to baseline. Seroconversion rates (defined as the percentage of participants with a post-vaccination titer of ≥4-fold) were 98% and 100%, respectively, compared to 1.3% for placebo.

Across the two age groups, anti-spike protein binding IgG GMTs were generally below the limit of quantification (200) at day 0 (Tables S7 and S9) reflecting the low rate of previous virus exposure in this population. Following first vaccination (day 21) of 5-µg (groups B and C combined) and 25-µg (groups D and E combined) NVX-CoV2373, anti-spike protein binding IgG GMTs were 1416 (95% CI, 1216 to 1649) and 3028 (95% CI, 2604 to 3522) for younger participants and 429 (95% CI, 360 to 512) and 959 (95% CI, 803 to 1146) for older participants; this equated to GMFRs of 12 and 25 relative to baseline for younger participants and 4 and 8 relative to baseline for older participants. Seroconversion rates were 79% and 93% for younger participants and 43% and 75% for older participants. Following second vaccination (day 35) of 5-µg (group B) and 25-µg (group D) NVX-CoV2373, anti-spike protein binding IgG GMTs increased to 65,019 (95% CI, 55,485 to 76,192) and 58,774 (95% CI, 51,612 to 66,930) for younger participants and 28,137 (95% CI, 21,617 to 36,623) and 32,871 (95% CI, 26,190 to 41,258) for older participants. This equated to GMFRs of 539 and 465 relative to baseline for younger participants and 258 and 286 relative to baseline for older participants. The approximate 50% decrement in the response for older participants was expected based on immunosenescence. Seroconversion rates also increased to 99% and 100% for younger participants and 97% and 99% for older participants.

#### Neutralizing Antibody Response

Across the two age groups in a subset of participants, neutralizing antibodies were generally below the limit of quantification (20) at day 0 (Tables S8, S10 and S11). Following first vaccination (day 21) of 5-µg (groups B and C combined) and 25-µg (groups D and E combined) NVX-CoV2373, neutralizing antibody GMTs were 41 (95% CI, 26 to 67) and 91 (95% CI, 48 to 175) for younger participants and 30 (95% CI, 16 to 54) and 33 (95% CI, 20 to 54) for older participants; this equated to GMFRs of 4 and 9 relative to baseline for younger participants and 3 and 3 relative to baseline for older participants. Seroconversion rates were 55% and 81% for younger participants and 39% and 50% for older participants. Following second vaccination (day 35) of 5-µg (group B) and 25-µg (group D) NVX-CoV2373, neutralizing antibody GMTs increased to 2201 (95% CI, 1343 to 3608) and 1783 (95% CI, 1192 to 2668) for younger participants and 981 (95% CI, 560 to 1717) and 1034 (95% CI, 640 to 1672) for older participants; this equated to GMFRs of 220 and 178 relative to baseline for younger participants and 98 and 103 relative to baseline for older participants. Seroconversion rates also increased to 100% and 100% for younger participants and 100% and 96% for older participants.

Neutralizing antibody responses of the two-dose regimens of 5-µg (group B) and 25-µg (group D) NVX-CoV2373 in both younger and older participants exceeded those seen in convalescent sera for both outpatient and hospitalized patients (Fig. 3).

**Figure 3.**
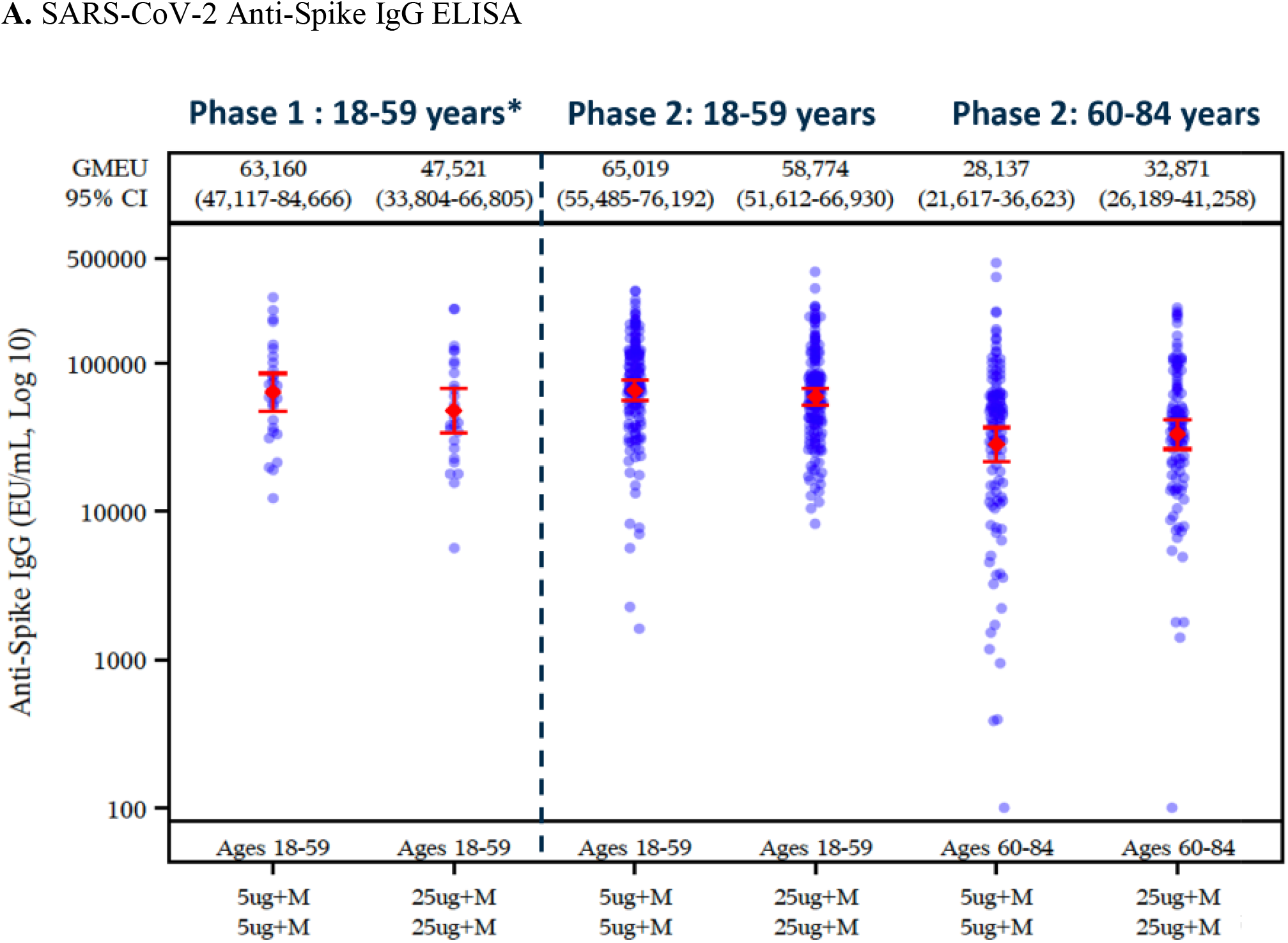
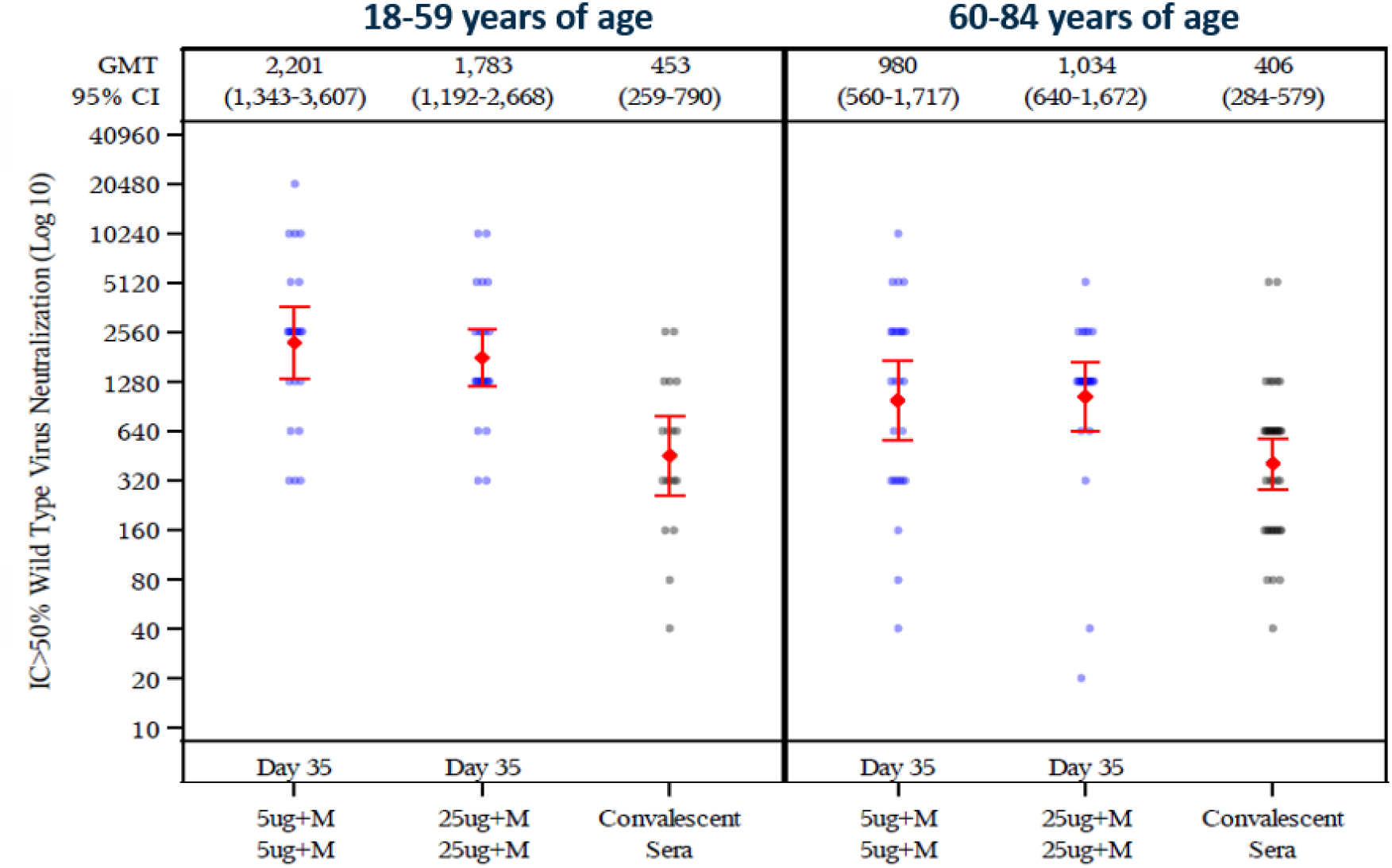
SARS-CoV-2 Anti-Spike IgG and Neutralizing Antibody Responses Through Day 35. Shown are geometric mean anti-spike IgG enzyme-linked immunosorbent assay (ELISA) unit responses to NVX-CoV2373 protein antigens (Panel A) for adult and older adult age groups in the phase 2 study for the two-dose primary vaccination treatment regimens (Groups B and D), along with aligned group results by the same assay in the phase 1 study, and wild-type SARS-CoV-2 microneutralization assay at an inhibitory concentration >50% (MN_50_) titer responses (Panel B) for adult and older adult age groups in the phase 2 study. In Panels A and B, boxes and horizontal bars represent interquartile range (IQR) and median area under the curve, respectively. Whisker endpoints are equal to the maximum and minimum values below or above the median ± 1.5 times the IQR.

## DISCUSSION

Based on the totality of the safety and immunogenicity data through day 35 from the phase 2 component of our phase 1-2 trial, the two-dose regimen of 5-µg NVX-CoV2373 administered 21 days apart was determined to be the optimal dose regimen for phase 3 development and licensure in both young and older adults. Following first vaccination, both dose levels of NVX-CoV2373 were well tolerated but there was a trend toward a higher incidence of local reactogenicity with the higher dose in both younger and older adults but no apparent differences in anti-spike protein binding IgG levels and neutralizing antibody responses by dose level. Following second vaccination, both dose levels of NVX-CoV2373 remained well tolerated despite increased frequencies and intensities of local and systemic reactogenicity in both younger and older adults, and a trend toward higher incidences of local and systemic reactogenicity with the higher dose remained. NVX-CoV2373 induced robust levels of anti-spike protein binding IgG levels and neutralizing antibodies, in both younger and older adults, with seroconversion rates of at least 96% achieved and no differences between the two dose levels. Notably, in both age groups, there was a high correlation between “binding” antibodies (anti-spike IgG) and neutralizing antibodies (Fig 4), suggesting the anti-spike immunity was predominantly functional, as indicated by neutralization. Based on the antibody responses, the low-dose, two-immunization regimen of 5-µg NVX-CoV2373 was selected to move into later phase development.

**Figure 4.**
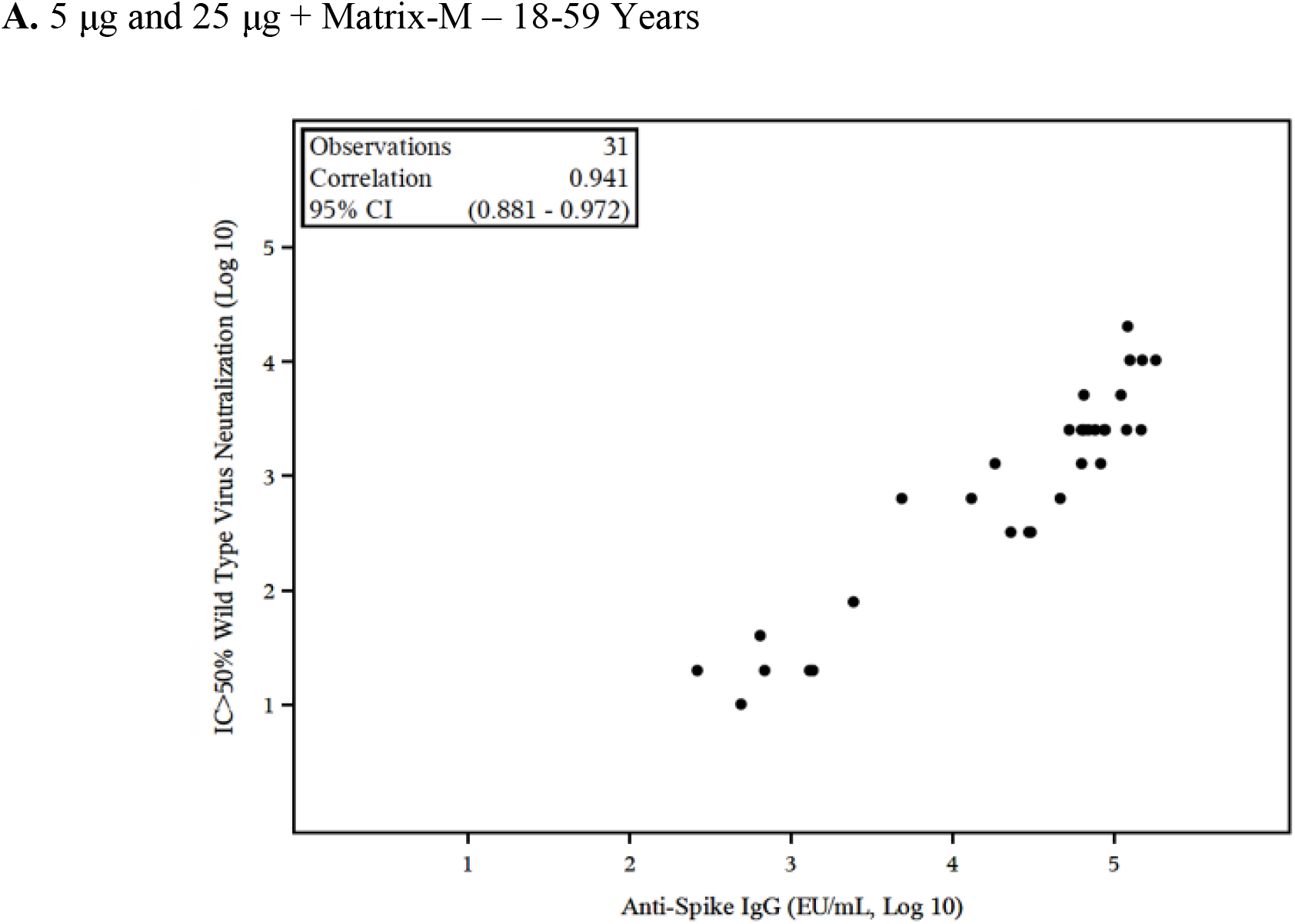
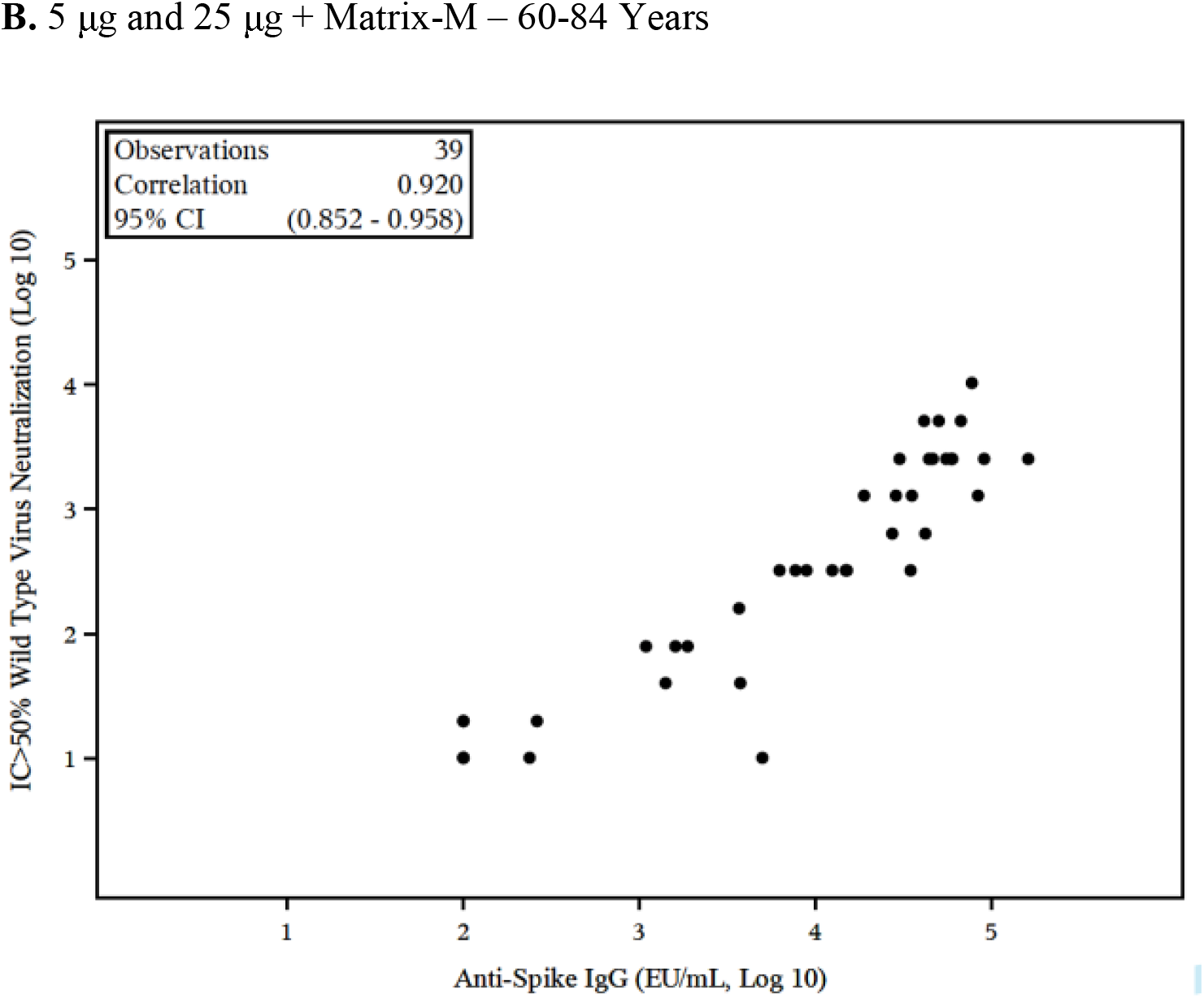
Correlation Between SARS-CoV-2 Anti-Spike IgG and Neutralizing Antibody Responses. Shown is a scattergram and correlation analysis of the neutralizing antibody titers and anti-spike IgG enzyme-linked immunosorbent assay (ELISA) units for the adjuvanted two-dose vaccine groups (Groups B and D) for adults 18 to 59 years (Panel A) and older adults 60 to 84 years (Panel B).

As expected, we found lower frequencies of local and systemic reactogenicity and lower anti-spike protein binding IgG and neutralizing antibody responses in older adults. These findings are consistent with other SARS-CoV-2 vaccines.^6,7^ Despite the lower immune response in older adults, seroconversion rates after the two-dose regimen of 5-µg NVX-CoV2373 were 97% for anti-spike protein binding IgG and 100% for neutralizing antibodies, which reflected increases from baseline of >250-fold and >95-fold, respectively. Additionally, in both younger and older adults, neutralizing antibody titers were similar to or exceeded those seen in a small sample of convalescent sera from hospitalized patients.

The safety and tolerability data from this study are also consistent with that gathered from over 14,000 participants, including over 4200 participants who received the Matrix-M1 adjuvant, enrolled in previous non-coronavirus nanoparticle trials. These studies included children, pregnant women, and older adults,^12-16^ with an age range from 5 months to 85 years of age and indicate that vaccines based on the nanoparticle/Matrix-M1 adjuvant technology have an acceptable safety profile to date.

Taken together, these data indicate that the Matrix-M1 adjuvanted rSARS-CoV-2 nanoparticle vaccine was highly immunogenic and well tolerated, with the trial largely reproducing the safety and immunogenicity from the phase 1 trial. Based on this, a phase 2a/b and two phase 3 studies evaluating the efficacy and safety of the two-dose regimen of 5-µg NVX-CoV2373 are ongoing in South Africa, the United Kingdom, the United States and Mexico.

## Data Availability

These are interim data, and individual participants remain masked to individual vaccine assignment. Therefore, it would be inappropriate to share individual level results at this time.

https://clinicaltrials.gov/ct2/show/NCT04368988?term=NVX-CoV2373+Covid-19+Vaccine&draw=2&rank=4

## [Acknowledgments]

We thank all of the study participants who volunteered for this study. Editorial assistance on the preparation of this manuscript was provided by Phase Five Communications, supported by Novavax, Inc.

Disclosure forms provided by the authors will be available with the full text of this article at publication

We acknowledge the contributions of the 2019nCoV-101 Study Group: Mark Adams, Mark Arya, Eugene Athan, Ira Berger, Paul Bradley, Richard Glover II, Paul Griffin, Joshua Kim, Scott Kitchener, Terry Klein, Amber Leah, Charlotte Lemech, Jason Lickliter, Mary Beth Manning, Fiona Napier-Flood, Paul Nugent, Susan Thackwray, and Mark Turner.

## References

1. World Health Organization. Coronavirus disease (COVID-19) dashboard. Geneva, Switzerland: World Health Organization. January 10 2021. https://covid19.who.int. Accessed February 18, 2021.

2. Bengtsson KL, Song H, Stertman L, et al. Matrix-M adjuvant enhances antibody, cellular and protective immune responses of a Zaire Ebola/Makona virus glycoprotein (GP) nanoparticle vaccine in mice. Vaccine 2016;34(16):1927–35.

3. Keech C, Albert G, Cho I, et al. Phase 1-2 trial of a SARS-CoV-2 recombinant spike protein nanoparticle vaccine. N Engl J Med 2020;383(24):2320–32.

4. Du L, He Y, Zhou Y, Liu S, Zheng BJ, Jiang S. The spike protein of SARS-CoV L a target for vaccine and therapeutic development. Nat Rev Microbiol 2009;7(3):226–36.

5. Tai W, He L, Zhang X, et al. Characterization of the receptor-binding domain (RBD) of 2019 novel coronavirus: implication for development of RBD protein as a viral attachment inhibitor and vaccine. Cell Mol Immunol 2020;17(6):613–20.

6. Baden LR, El Sahly HM, Essink B, et al; COVE Study Group. Efficacy and safety of the mRNA-1273 SARS-CoV-2 vaccine. N Engl J Med 2021;384(5):403–16. Epub 2020 Dec 30.

7. Polack FP, Thomas SJ, Kitchin N, et al; C4591001 Clinical Trial Group. Safety and efficacy of the BNT162b2 mRNA Covid-19 vaccine. N Engl J Med 2020;383(27):2603–15. Epub 2020 Dec 10.

8. Karthik K, Senthilkumar TMA, Udhayavel S, Raj GD. Role of antibody-dependent enhancement (ADE) in the virulence of SARS-CoV-2 and its mitigation strategies for the development of vaccines and immunotherapies to counter COVID-19. Hum Vaccin Immunother 2020;16(12):3055–60.

9. Food and Drug Administration. Guidance for Industry: Toxicity Grading Scale for Healthy Adult and Adolescent Volunteers Enrolled in Preventive Vaccine Clinical Trials. September 2007. https://www.fda.gov/regulatory-information/search-fda-guidance-documents/toxicity-grading-scale-healthy-adult-and-adolescent-volunteers-enrolled-preventive-vaccine-clinical. Accessed February 17, 2021.

10. Division of AIDS (DAIDS), National Institutes of Health. Division of AIDS (DAIDS) Table for Grading the Severity of Adult and Pediatric Adverse Events. Version 2.1. July 2017. https://rsc.niaid.nih.gov/sites/default/files/daidsgradingcorrectedv21.pdf. Accessed February 17, 2021.

11. D2.3 Priority List of Adverse Events of Special Interest: COVID-19. V2.0. May 25, 2020. https://brightoncollaboration.us/wp-content/uploads/2020/06/SPEAC_D2.3_V2.0_COVID-19_20200525_public.pdf. Accessed June 19, 2020.

12. Fries L, Shinde V, Stoddard JJ, et al. Immunogenicity and safety of a respiratory syncytial virus fusion protein (RSV F) nanoparticle vaccine in older adults. Immun Ageing 2017;14:8.

13. Munoz FM, Swamy GK, Hickman SP, et al. Safety and immunogenicity of a respiratory syncytial virus fusion (F) protein nanoparticle vaccine in healthy third-trimester pregnant women and their infants. J Infect Dis 2019;220:1802–15.

14. Glenn GM, Fries LF, Thomas DN, et al. A randomized, blinded, controlled, dose-ranging study of a respiratory syncytial virus recombinant fusion (F) nanoparticle vaccine in healthy women of childbearing age. J Infect Dis 2016;213:411–22.

15. Shinde V, Cai R, Plested J, et al. Induction of cross-reactive hemagglutination inhibiting antibody and polyfunctional CD4+ T-cell responses by a recombinant Matrix-M-adjuvanted hemagglutinin nanoparticle influenza vaccine. medRxiv 2020:2020.05.11.20098574.

16. Shinde V, Cho I, Plested JS, et al. Comparison of the safety and immunogenicity of a novel Matrix-M-adjuvanted nanoparticle influenza vaccine with a quadrivalent seasonal influenza vaccine in older adults: a randomized controlled trial. medRxiv 2020.08.07.20170514; doi: https://doi.org/10.1101/2020.08.07.20170514.

