## Supplemental Text, Figures and Tables for "Evaluation of a SARS-CoV-2 Vaccine NVX-CoV2373 in Younger and Older Adults"

This appendix has been provided by the authors to give readers additional information about their work.

**2019nCoV-101 Study Group Members**

**(listed in PubMed, and ordered alphabetically by institutional affiliation)**

The following study group members were all closely involved with the design, implementation, and oversight of the phase 2 part of the NVX-CoV2373-2019nCoV-101 clinical trial.

Advanced Clinical Research, Meridian, Idaho, United States:

Alliance for Multispecialty Research, Wichita and Newton, Kansas, United States:

Australian Clinical Research Network, Maroubra, NSW, Australia:

Barwon Health, Geelong, Melbourne, Victoria, Australia:

Central Kentucky Research Associates, Lexington, Kentucky, United States:

Meridian Clinical Research, Rockville, Maryland, and Savannah, Georgia, United States:

Novavax, Inc., Gaithersburg, Maryland, United States:

Nucleus Network Pty Ltd, Melbourne, Victoria, and Herston, Queensland, Australia:

Paratus Clinical Research – Canberra, ACT, and Central Coast and Western Sydney, NSW, Australia:

Rapid Medical Research, Cleveland, Ohio, United States:

Scientia Clinical Research Limited, Randwick, NSW, Australia:

Synexus Clinical Research US, Cincinnati, Ohio, United States:

University of the Sunshine Coast, Sippy Downs and Morayfield, Queensland, Australia:

### Supplemental Methods:

#### Vaccination Pause Rules

Adverse events meeting any one of the following criteria will result in an immediate enrollment or further dosing hold for Part 2 of the study, pending further review by the Safety Monitoring Committee (SMC) at the direction of the SMC Chair:

- Any toxicity grade 3 (severe) solicited single adverse event term across all study vaccine groups (blinded) occurring in ≥5% of participants for either study age group (after a minimum of 100 participants are enrolled in either age group) following vaccination (first, second vaccinations and booster dose to be assessed separately).
- Any grade 3 (severe) unsolicited single adverse event preferred term for which the investigator assesses as related which occurs in ≥7 participants overall (both age groups) within 35 days following first dose vaccination.

In addition, any serious adverse event assessed as related to vaccine (by study investigator and/or the sponsor) will be reported by the sponsor to the SMC Chair as soon as possible, and within 24 hours of the sponsor’s awareness of the event. Based on this initial report of the event to the SMC Chair, the Chair may advise the sponsor to immediately pause enrollment and further dosing in either some or all participants in the study and to convene an ad hoc meeting, or make alternative recommendations. The SMC Charter defines processes for how this review will occur and how the Chair’s recommendations will be documented.

The sponsor, along with medical monitor, may request an SMC review for any safety concerns that may arise in the trial and not associated with any specific pause rule.

#### Immunologic Assay Method Details

SARS-CoV-2 Spike Protein Serum IgG ELISA. SARS-CoV-2 rS vaccine protein (NVX-CoV2373) was immobilized onto the surface of the 96-well microtiter plate wells (100 µL per well) by direct adsorption for 15 to 96 hours at 2°C to 8°C at a concentration of 1 µg/mL in PBS. Plates were washed 3 times with 300 µL/well PBST, blocked with 300 µL blocking buffer for 1 to 1.5 hours at 24°C ± 2°C. Diluted reference standard (2-fold dilution series of 12 dilutions starting 1:1000) and human serum samples (3-fold dilution series of 12 dilutions) in assay buffer (1% milk in PBS) starting at 1:100 dilution are then added in duplicate (100 µL per well) to the SARS-CoV-2 rS vaccine protein-coated wells and any specific antibodies are allowed to complex with the coated antigen for 2 hours at 24°C ± 2°C. Plates are washed 2 times with 300 µL/well PBST. Antibodies bound to the SARS-CoV-2 rS vaccine protein are then detected using a horseradish peroxidase (HRP) conjugate goat anti-human IgG antibody diluted 1:2000 (Southern Biotech cat no. 2040-05) incubated for 1 hour ± 10 minutes at 24°C ± 2°C, washed 3 times with 300 µL/well PBST and a colorimetric signal generated by addition of 100 µL per well 3, 3′,5,5′-tetramethylbenzidine (TMB) chromogenic substrate for 10 minutes ± 2 minutes at 24°C ± 2°C. After incubation was complete, the TMB reaction was stopped with 100 µL per well of TMB Stop solution. The absorbance was measured at 450 nm using a molecular device 96-well plate reader. When binding reagents (coated antigen and secondary antibody) are in excess, the optical density (OD) of the chromogenic substrate at endpoint is proportional to the quantity of anti-SARS-CoV-2 rS IgG present in the serum sample. The total anti-SARS-CoV-2 S protein IgG antibody level in a serum sample was quantitated in ELISA unit, EU/mL by comparison to a reference standard curve. The results were analyzed by SoftMax Pro software using 4-PL curve fit. Assay included control plates comprising of positive controls and negative control.

SARS-CoV-2 Microneutralization Assay. The viral working stock was SARS-CoV-2 hCoV-19/Australia/VIC01/2020 (GenBank MT007544.1) was a gift from Melbourne’s Peter Doherty Institute for Infection and Immunity (Melbourne, Australia). Sequence analysis of the SARS-CoV-2 hCoV-19/Australia/VIC01/2020 spike protein shows high sequence homology with the Wuhan strain with only one nucleotide difference in the spike protein (S247R) compared to the Wuhan strain. All serum samples were heat inactivated at 56°C for 30 minutes to remove complement and allowed to equilibrate to room temperature prior to processing for neutralization titer. Sera were diluted by addition of 28 µL to 252 µL of assay media in a 96-well dilution plate and an 11-point 2-fold serial dilution was prepared. Sera were then mixed with an equal volume (140 µL) of SARS-CoV-2 (4000 TCID50 units/mL) and incubated for 1 hour at 37^o^C, 5% CO_2_. The MN assay media (AM) comprised Dulbecco Essential Medium without L-glutamine (DMEM; Thermo Fisher Scientific, cat. no. 10313-021) supplemented with 2% FBS (Bovagen SFBS), 1% Glutamax (Thermo Fisher Scientific, cat. no. 35050-061), and 1% Pen/Strep (Thermo Fisher Scientific, cat. no. 15140-122).

Following this incubation, 100 µL of the virus/serum mixtures (100 TCID50 units/well) was added in duplicate to Vero E6 cells, pre-seeded 24 hours prior in 96-well plates in 100 µL of assay media at 1.5 x 104 cells/well plates were incubated for 3 days at 37^o^C, 5% CO_2_.

Following this incubation, 100 µL of the virus/serum mixtures (100 TCID50 units/well) was added in duplicate to Vero E6 cells, pre-seeded 24 hours prior in 96-well plates in 100 µL of assay media at 1.5 x 104 cells/well plates were incubated for 3 days at 37^o^C, 5% CO_2_.

The residual non-neutralized virus is detected via cytopathic effect (CPE) by microscopic scoring. Two replicate wells per dilution are scored as either positive (SARS-CoV-2 cytopathology is present) or negative (healthy Vero E6 monolayer). The neutralization titer is expressed as the reciprocal of the highest dilution at which 50% of the replicate wells are protected from infection (MN50).

The final serum dilution range of the assay was MN50 titer range of 20‒20,480. If at least 50% protection was not observed for an individual serum at any dilution, the MN50 titer was recorded as <20. Not less than 4 virus control only wells, and mock infected wells (no virus) were run per plate.

##### Table S1. Toxicity Grading Scales for Solicited Local and Systemic Adverse Events – Modified From FDA Toxicity Grading Scale for Clinical Abnormalities.

| Local Reaction to Injectable Product | Mild (Grade 1) | Moderate  (Grade 2) | Severe (Grade 3) | Potentially Life Threatening  (Grade 4) |
| --- | --- | --- | --- | --- |
| Pain | Does not interfere with activity | Repeated use of non-prescription pain reliever >24 hours or interferes with activity | Significant; any use of prescription pain reliever or prevents daily activity | Requires ER visit or hospitalization |
| Tenderness | Mild discomfort to touch | Discomfort with movement | Significant discomfort at rest | Requires ER visit or hospitalization |
| Erythema/redness* | 2.5 − 5 cm | 5.1 − 10 cm | >10 cm | Necrosis or exfoliative dermatitis† |
| Induration/swelling* | 2.5 − 5 cm | 5.1 − 10 cm | >10 cm | Necrosis† |
| **Systemic (General)** | | | | |
|  | **Mild (Grade 1)** | **Moderate (Grade 2)** | **Severe (Grade 3)** | **Potentially Life Threatening (Grade 4)** |
| Fever‡ (°C)  (°F) | 38.0 − 38.4 100.4 − 101.1 | 38.5 − 38.9 101.2 − 102.0 | 39.0 − 40 102.1 − 104 | >40 >104 |
| Nausea/vomiting | Does not interfere with activity or 1 − 2 episodes/24 hours | Some interference with activity or >2 episodes/24 hours | Prevents daily activity, or requires IV hydration outside of hospital | Requires ER visit or hospitalization |
| Headache | Does not interfere with activity | Repeated use of non-prescription pain reliever >24 hours or interferes with activity | Significant; any use of prescription pain reliever or prevents daily activity | Requires ER visit or hospitalization |
| Fatigue/Malaise | Does not interfere with activity | Some interference with activity | Significant, prevents daily activity | Requires ER visit or hospitalization |
| Myalgia | Does not interfere with activity | Some interference with activity | Significant, prevents daily activity | Requires ER visit or hospitalization |
| Arthralgia | Does not interfere with activity | Some interference with activity | Significant, prevents daily activity | Requires ER visit or hospitalization |
| * The measurements should be recorded as a continuous variable.  † These events are not subject reported through the electronic diary and will be monitored through the adverse event pages of the study database.  ‡ Oral temperature if subject collected, sites may collect temperature using local clinic practices/devices. Toxicity grade will be derived. | | | | |

| Table S2. Adverse Events of Special Interest Relevant to COVID-19.* | |
| --- | --- |
| **Body System** | **Diagnoses** |
| Immunologic | Enhanced disease following immunization, cytokine release syndrome related to COVID-19 disease,† multisystem inflammatory syndrome in children (MIS-C) |
| Respiratory | Acute respiratory distress syndrome (ARDS) |
| Cardiac | Acute cardiac injury including:   - Microangiopathy - Heart failure and cardiogenic shock - Stress cardiomyopathy - Coronary artery disease - Arrhythmia - Myocarditis, pericarditis |
| Hematologic | Coagulation disorder   - Deep vein thrombosis - Pulmonary embolus - Cerebrovascular stroke - Limb ischemia - Hemorrhagic disease - Thrombotic complications |
| Renal | Acute kidney injury |
| Gastrointestinal | Liver injury |
| Neurologic | Guillain Barré Syndrome, anosmia, ageusia, meningoencephalitis |
| Dermatologic | Chilblain-like lesions, single organ cutaneous vasculitis, erythema multiforme |
| * COVID-19 disease manifestations associated with more severe presentation and decompensation with consideration of enhanced disease potential. The current listing is based on Safety Platform for Emergency Vaccines (SPEAC) D2.3 Priority List of Adverse Events of Special Interest: COVID-19 (SPEAC 2020).  † Cytokine release syndrome related to COVID-19 disease is a disorder characterized by nausea, headache, tachycardia, hypotension, rash, and/or shortness of breath. | |

| Table S3. Potential Immune-Mediated Medical Conditions. | |
| --- | --- |
| **Categories** | **Diagnoses (as MedDRA Preferred Terms)** |
| Neuroinflammatory Disorders: | Acute disseminated encephalomyelitis (including site specific variants: eg, non-infectious encephalitis, encephalomyelitis, myelitis, myeloradiculomyelitis), cranial nerve disorders including paralyses/paresis (eg, Bell’s palsy), generalized convulsion, Guillain-Barre syndrome (including Miller Fisher syndrome and other variants), immune mediated peripheral neuropathies and plexopathies (including chronic inflammatory demyelinating polyneuropathy, multifocal motor neuropathy and polyneuropathies associated with monoclonal gammopathy), myasthenia gravis, multiple sclerosis, narcolepsy, optic neuritis, transverse myelitis, uveitis |
| Musculoskeletal and Connective Tissue Disorders: | Antisynthetase syndrome, dermatomyositis, juvenile chronic arthritis (including Still’s disease), mixed connective tissue disorder, polymyalgia rheumatic, polymyositis, psoriatic arthropathy, relapsing polychondritis, rheumatoid arthritis, scleroderma (including diffuse systemic form and CREST syndrome), spondyloarthritis (including ankylosing spondylitis, reactive arthritis [Reiter's Syndrome] and undifferentiated spondyloarthritis), systemic lupus erythematosus, systemic sclerosis, Sjogren’s syndrome |
| Vasculitides: | Large vessels vasculitis (including giant cell arteritis such as Takayasu's arteritis and temporal arteritis), medium sized and/or small vessels vasculitis (including polyarteritis nodosa, Kawasaki's disease, microscopic polyangiitis, Wegener's granulomatosis, Churg–Strauss syndrome [allergic granulomatous angiitis], Buerger’s disease [thromboangiitis obliterans], necrotizing vasculitis and anti-neutrophil cytoplasmic antibody [ANCA] positive vasculitis [type unspecified], Henoch-Schonlein purpura, Behcet's syndrome, leukocytoclastic vasculitis) |
| Gastrointestinal Disorders: | Crohn’s disease, celiac disease, ulcerative colitis, ulcerative proctitis |
| Hepatic Disorders: | Autoimmune hepatitis, autoimmune cholangitis, primary sclerosing cholangitis, primary biliary cirrhosis |
| Renal Disorders: | Autoimmune glomerulonephritis (including IgA nephropathy, glomerulonephritis rapidly progressive, membranous glomerulonephritis, membranoproliferative glomerulonephritis, and mesangioproliferative glomerulonephritis |
| Cardiac Disorders: | Autoimmune myocarditis/cardiomyopathy |
| Skin Disorders: | Alopecia areata, psoriasis, vitiligo, Raynaud’s phenomenon, erythema nodosum, autoimmune bullous skin diseases (including pemphigus, pemphigoid and dermatitis herpetiformis), cutaneous lupus erythematosus, morphoea, lichen planus, Stevens-Johnson syndrome, Sweet’s syndrome |
| Hematologic Disorders: | Autoimmune hemolytic anemia, autoimmune thrombocytopenia, antiphospholipid syndrome, thrombocytopenia |
| Metabolic Disorders: | Autoimmune thyroiditis, Grave’s or Basedow’s disease, Hashimoto thyroiditis^*^, diabetes mellitus type 1, Addison’s disease |
| Other Disorders: | Goodpasture syndrome, idiopathic pulmonary fibrosis, pernicious anemia, sarcoidosis |
| * For Hashimoto thyroiditis: new onset only. | |

### Supplemental Figure and Tables

**Figure S1. Vaccine Regimens and Key Trial Assessments.**

Shown are the planned randomization schema and associated vaccine regimens administered in the trial (Panel A), along with timing of the key safety and immunogenicity assessments (Panel B).

| **A: Vaccine Regimens** | | | | | | | | | | | |
| --- | --- | --- | --- | --- | --- | --- | --- | --- | --- | --- | --- |
| **Vaccine**  **Group** | **No. of Participants** | | **Day 0** | | **Day 21**  **–1 to +3 days** | |  | **Vaccine Group** | **Day 189**  **±15 days** | |  |
|  | Randomized | | rSARS-CoV-2 | Matrix-M1  adjuvant | rSARS-CoV-2 | Matrix-M1  adjuvant |  |  | rSARS-CoV-2 | Matrix-M1  adjuvant |  |
| A | 255 | | 0 | 0 | 0 | 0 |  | A | 0 | 0 |  |
| B | 257 | | 5 μg | 50 μg | 5 μg | 50 μg |  | B1  B | 0  5 μg | 0  50 μg |  |
| C | 257 | | 5 μg | 50 μg | 5 μg | 0 |  | C1  C2 | 0  5 μg | 0  50 μg |  |
| D | 258 | | 25 μg | 50 μg | 25 μg | 50 μg |  | D | 0 | 0 |  |
| E | 256 | | 25 μg | 50 μg | 25 μg | 0 |  | E | 0 | 0 |  |
| **B: Key Trial Timings** | | | | | | | | | | | |
| **Procedure** | | **Screening** | **Day** | | | | | | | | |
|  | | -45 to Day 0 | 0 | 7 | 21 | 28 | 35 | 105 | 189 | 217 | 357 |
| Vaccination | |  | **X** |  | **X** |  |  |  | **X** |  |  |
| Blood sample: immunogenicity (serology) | | X | **X** |  | **X** |  | **X** |  | **X** | **X** | **X** |
| Blood sample: immunogenicity (CMI subset) | |  |  | **X** |  | **X** |  |  |  |  |  |
| Nasal swab | | **X** |  | | | | | | | | |
| monitoring for COVID-19 disease endpoints | |  |  |  |  |  | | | | | |
| Reactogenicity – 7 days after each dose | |  |  | |  | |  |  |  |  |  |
| Unsolicited adverse event∞ | |  |  | | | | |  |  |  |  |
| Medically attended adverse event: All/related | |  |  | | | | | | | |  |
| Serious adverse event or adverse event of special interest | |  |  | | | | | | | | |

##### Table S4a. Percentage of All Participants Experiencing Solicited Local and Systemic Adverse Events by Symptom, Vaccination Dose, Vaccine Group, and Maximum Toxicity Grade (Safety Analysis Set).*

| **Symptom** | **Vaccination Dose** | **Vaccine Group†** | | **N** | **% Grade 0**  **(None)** | **% Grade 1**  **(Mild)** | **% Grade 2**  **(Moderate)** | **% Grade 3**  **(Severe)** | **% Grade 4**  **(Potentially Life-Threatening)** |
| --- | --- | --- | --- | --- | --- | --- | --- | --- | --- |
| Any solicited local AE | 1 | A  B  C  D  E | 0/0  5/50  5/50  25/50  25/50 | 252  253  255  252  253 | 84.5  48.2  47.1  38.9  30.8 | 14.3  37.2  36.9  39.3  43.5 | 1.2  14.2  16.1  21.0  25.3 | 0  0.4  0  0.8  0.4 | 0  0  0  0  0 |
|  | 2 | A  B  C  D  E | 0/0  5/50  0/0  25/50  0/0 | 242  250  249  247  236 | 90.9  30.0  89.2  19.0  91.5 | 8.3  38.8  8.0  38.5  7.6 | 0.8  26.0  2.8  34.0  0.8 | 0  5.2  0  7.7  0 | 0  0  0  0.8  0 |
| Pain | 1 | A  B  C  D  E | 0/0  5/50  5/50  25/50  25/50 | 252  253  255  252  253 | 96.0  73.1  72.2  67.1  58.5 | 4.0  25.7  27.5  32.1  38.3 | 0  1.2  0.4  0.8  3.2 | 0  0  0  0  0 | 0  0  0  0  0 |
|  | 2 | A  B  C  D  E | 0/0  5/50  0/0  25/50  0/0 | 242  250  249  247  236 | 96.3  54.4  93.6  45.3  94.9 | 3.7  39.6  6.0  42.9  5.1 | 0  4.0  0.4  9.3  0 | 0  2.0  0  2.0  0 | 0  0  0  0.4  0 |
| Erythema | 1 | A  B  C  D  E | 0/0  5/50  5/50  25/50  25/50 | 252  253  255  252  253 | 100.0  99.2  99.6  99.6  99.2 | 0  0.8  0.4  0  0 | 0  0  0  0.4  0.8 | 0  0  0  0  0 | 0  0  0  0  0 |
|  | 2 | A  B  C  D  E | 0/0  5/50  0/0  25/50  0/0 | 242  250  249  247  236 | 100.0  95.2  100.0  86.6  100.0 | 0  1.6  0  2.8  0 | 0  2.0  0  7.3  0 | 0  1.2  0  3.2  0 | 0  0  0  0  0 |
| Swelling | 1 | A  B  C  D  E | 0/0  5/50  5/50  25/50  25/50 | 252  253  255  252  253 | 99.6  99.2  98.8  99.6  98.8 | 0.4  0.8  0  0.4  0.4 | 0  0  1.2  0  0.8 | 0  0  0  0  0 | 0  0  0  0  0 |
|  | 2 | A  B  C  D  E | 0/0  5/50  0/0  25/50  0/0 | 242  250  249  247  236 | 100.0  94.4  100.0  89.1  100.0 | 0  2.8  0  4.0  0 | 0  2.4  0  4.9  0 | 0  0.4  0  2.0  0 | 0  0  0  0  0 |
| Tenderness | 1 | A  B  C  D  E | 0/0  5/50  5/50  25/50  25/50 | 252  253  255  252  253 | 86.9  51.8  52.2  43.7  37.5 | 11.9  34.0  32.5  35.3  39.1 | 1.2  13.8  15.3  20.2  22.9 | 0  0.4  0  0.8  0.4 | 0  0  0  0  0 |
|  | 2 | A  B  C  D  E | 0/0  5/50  0/0  25/50  0/0 | 242  250  249  247  236 | 92.6  34.8  91.2  23.9  92.8 | 6.6  36.4  6.4  41.3  6.4 | 0.8  25.2  2.4  29.6  0.8 | 0  3.6  0  4.5  0 | 0  0  0  0.8  0 |
| Any solicited systemic AE | 1 | A  B  C  D  E | 0/0  5/50  5/50  25/50  25/50 | 251  255  255  252  253 | 63.7  56.1  60.00  55.6  60.5 | 20.7  25.5  23.5  30.2  23.3 | 13.9  14.5  15.3  13.1  14.6 | 0.8  3.9  1.2  1.2  1.2 | 0.8  0  0  0  0.4 |
|  | 2 | A  B  C  D  E | 0/0  5/50  0/0  25/50  0/0 | 241  250  249  247  235 | 72.6  47.2  71.5  36.4  78.3 | 15.8  22.4  17.3  24.3  13.6 | 10.4  24.8  9.6  30.8  8.1 | 0.8  5.6  1.6  8.1  0 | 0.4  0  0  0.4  0 |
| Joint pain/ arthralgia | 1 | A  B  C  D  E | 0/0  5/50  5/50  25/50  25/50 | 251  255  255  252  253 | 94.0  93.3  91.8  95.2  93.7 | 4.4  3.1  4.3  4.0  3.2 | 1.6  2.7  3.9  0.8  2.8 | 0  0.8  0  0  0.4 | 0  0  0  0  0 |
|  | 2 | A  B  C  D  E | 0/0  5/50  0/0  25/50  0/0 | 241  250  249  247  235 | 96.3  85.2  96.8  81.0  98.3 | 2.1  7.2  1.6  6.1  0.9 | 1.7  6.4  1.6  11.3  0.9 | 0  1.2  0  1.6  0 | 0  0  0  0  0 |
| Fatigue | 1 | A  B  C  D  E | 0/0  5/50  5/50  25/50  25/50 | 251  255  255  252  253 | 79.3  76.9  75.7  83.7  81.4 | 10.8  11.4  13.3  9.1  11.1 | 9.6  9.8  9.8  6.3  7.1 | 0.4  2.0  1.2  0.8  0.4 | 0  0  0  0  0 |
|  | 2 | A  B  C  D  E | 0/0  5/50  0/0  25/50  0/0 | 241  250  249  247  235 | 86.3  64.4  82.3  57.5  96.8 | 6.6  12.8  9.6  12.6  7.7 | 6.6  20.0  7.2  23.5  5.5 | 0.4  2.8  0.8  6.5  0 | 0  0  0  0  0 |
| Malaise | 1 | A  B  C  D  E | 0/0  5/50  5/50  25/50  25/50 | 251  255  255  252  253 | 88.0  87.8  87.8  90.9  89.7 | 6.4  6.3  6.7  5.6  7.1 | 5.2  3.5  4.7  2.8  3.2 | 0  2.4  0.8  0.8  0 | 0.4  0  0  0  0 |
|  | 2 | A  B  C  D  E | 0/0  5/50  0/0  25/50  0/0 | 241  250  249  247  235 | 92.1  73.6  92.4  70.0  95.3 | 5.0  9.2  4.0  7.7  3.8 | 2.9  14.8  3.2  17.8  0.9 | 0  2.4  0.4  4.5  0 | 0  0  0  0  0 |
| Temperature/ Fever | 1 | A  B  C  D  E | 0/0  5/50  5/50  25/50  25/50 | 248  255  255  252  253 | 97.6  97.6  97.6  98.8  98.4 | 0.8  1.2  1.6  0  0.4 | 1.2  0  0.8  0.8  0.8 | 0  1.2  0  0.4  0 | 0.4  0  0  0  0.4 |
|  | 2 | A  B  C  D  E | 0/0  5/50  0/0  25/50  0/0 | 239  249  249  245  230 | 99.2  95.6  99.6  91.8  99.6 | 0.4  3.2  0  3.7  0 | 0  0.8  0.4  3.7  0.4 | 0  0.4  0  0.8  0 | 0.4  0  0  0  0 |
| Headache | 1 | A  B  C  D  E | 0/0  5/50  5/50  25/50  25/50 | 251  255  255  252  253 | 79.3  76.9  75.7  83.7  81.4 | 10.8  11.4  13.3  9.1  11.1 | 9.6  9.8  9.8  6.3  7.1 | 0.4  2.0  1.2  0.8  0.4 | 0  0  0  0  0 |
|  | 2 | A  B  C  D  E | 0/0  5/50  0/0  25/50  0/0 | 241  250  249  247  235 | 87.1  70.4  87.1  66.0  88.1 | 10.4  16.8  10.4  16.6  10.2 | 2.1  10.8  2.0  15.0  1.7 | 0.4  2.0  0.4  1.4  0 | 0  0  0  0  0 |
| Muscle pain/ Myalgia | 1 | A  B  C  D  E | 0/0  5/50  5/50  25/50  25/50 | 251  255  255  252  253 | 97.4  98.3  99.1  99.1  97.8 | 2.2  1.3  0.9  0.9  2.2 | 0.4  0.4  0  0  0 | 0  0  0  0  0 | 0  0  0  0  0 |
|  | 2 | A  B  C  D  E | 0/0  5/50  0/0  25/50  0/0 | 241  250  249  247  235 | 93.4  69.2  92.8  59.1  97.9 | 4.6  13.6  4.8  18.6  1.3 | 2.1  14.8  2.0  18.6  0.9 | 0  2.4  0.4  3.2  0 | 0  0  0  0.4  0 |
| Nausea or vomiting | 1 | A  B  C  D  E | 0/0  5/50  5/50  25/50  25/50 | 251  255  255  252  253 | 99.6  99.1  99.6  99.6  98.7 | 0.4  0.4  0.4  0.4  1.3 | 0  0.4  0  0  0 | 0  0  0  0  0 | 0  0  0  0  0 |
|  | 2 | A  B  C  D  E | 0/0  5/50  0/0  25/50  0/0 | 241  250  249  247  235 | 96.3  92.8  98.0  89.1  97.4 | 3.7  6.0  1.6  6.1  2.1 | 0  1.2  0.4  4.5  0.4 | 0  0  0  0.4  0 | 0  0  0  0  0 |

*	AE denotes adverse event, , N denotes number of participants assessed.

†	Group A (placebo): 0 µg NVX-CoV2373/0 µg Matrix-M1 on first vaccination; 0 µg NVX-CoV2373/0 µg Matrix-M1 on second vaccination. Group B: 5 µg NVX-CoV2373/50 µg Matrix-M1 on first vaccination; 5 µg NVX-CoV2373/50 µg Matrix-M1 on second vaccination. Group C: 5 µg NVX-CoV2373/50 µg Matrix-M1 on first vaccination; 0 µg NVX-CoV2373/0 µg Matrix-M1 on second vaccination. Group D: 25 µg NVX-CoV2373/50 µg Matrix-M1 on first vaccination; 25 µg NVX-CoV2373/50 µg Matrix-M1 on second vaccination. Group E: 25 µg NVX-CoV2373/50 µg Matrix-M1 on first vaccination; 0 µg NVX-CoV2373/0 µg Matrix-M1 on second vaccination.

##### Table S4b. Percentage of Older Adults (60 to 84 Years) Experiencing Solicited Local and Systemic Adverse Events by Symptom, Vaccination Dose, Vaccine Group, and Maximum Toxicity Grade (Safety Analysis Set).*

| **Symptom** | **Vaccination Dose** | **Vaccine Group†** | | **N** | **% Grade 0**  **(None)** | **% Grade 1**  **(Mild)** | **% Grade 2**  **(Moderate)** | **% Grade 3**  **(Severe)** | **% Grade 4**  **(Potentially Life-Threatening)** |
| --- | --- | --- | --- | --- | --- | --- | --- | --- | --- |
| Any solicited local AE | 1 | A  B  C  D  E | 0/0  5/50  5/50  25/50  25/50 | 114  114  117  113  117 | 87.7  61.4  63.2  55.8  40.2 | 11.4  36.0  32.5  38.1  43.6 | 0.9  2.6  4.3  6.2  15.4 | 0  0  0  0  0.9 | 0  0  0  0  0 |
|  | 2 | A  B  C  D  E | 0/0  5/50  0/0  25/50  0/0 | 110  113  113  110  109 | 93.6  38.1  92.9  32.7  89.9 | 5.5  42.5  5.3  39.1  10.1 | 0.9  16.8  1.8  19.1  0 | 0  2.7  0  8.2  0 | 0  0  0  0.9  0 |
| Pain | 1 | A  B  C  D  E | 0/0  5/50  5/50  25/50  25/50 | 114  114  117  113  117 | 98.2  82.5  82.9  81.4  69.2 | 1.8  16.7  17.1  18.6  30.8 | 0  0.9  0  0  0 | 0  0  0  0  0 | 0  0  0  0  0 |
|  | 2 | A  B  C  D  E | 0/0  5/50  0/0  25/50  0/0 | 110  113  113  110  109 | 97.3  59.3  95.6  60.9  94.5 | 2.7  39.8  4.4  30.9  5.5 | 0  0  0  7.3  0 | 0  0.9  0  0  0 | 0  0  0  0.9  0 |
| Erythema | 1 | A  B  C  D  E | 0/0  5/50  5/50  25/50  25/50 | 114  114  117  113  117 | 100.0  100.0  99.1  99.1  99.1 | 0  0  0.9  0  0 | 0  0  0  0.9  0.9 | 0  0  0  0  0 | 0  0  0  0  0 |
|  | 2 | A  B  C  D  E | 0/0  5/50  0/0  25/50  0/0 | 110  113  113  110  109 | 100.0  94.7  100.0  84.5  100.0 | 0  1.8  0  2.7  0 | 0  2.7  0  7.3  0 | 0  0.9  0  5.5  0 | 0  0  0  0  0 |
| Swelling | 1 | A  B  C  D  E | 0/0  5/50  5/50  25/50  25/50 | 114  114  117  113  117 | 100.0  100.0  99.1  100.0  97.4 | 0  0  0  0  0.9 | 0  0  0.9  0  1.7 | 0  0  0  0  0 | 0  0  0  0  0 |
|  | 2 | A  B  C  D  E | 0/0  5/50  0/0  25/50  0/0 | 110  113  113  110  109 | 100.0  92.0  100.0  90.0  100.0 | 0  3.5  0  2.7  0 | 0  4.4  0  4.5  0 | 0  0  0  2.7  0 | 0  0  0  0  0 |
| Tenderness | 1 | A  B  C  D  E | 0/0  5/50  5/50  25/50  25/50 | 114  114  117  113  117 | 89.5  65.8  68.4  59.3  43.6 | 9.6  31.6  27.4  35.4  41.9 | 0.9  2.6  4.3  5.3  13.7 | 0  0  0  0  0.9 | 0  0  0  0  0 |
|  | 2 | A  B  C  D  E | 0/0  5/50  0/0  25/50  0/0 | 110  113  113  110  109 | 94.5  45.1  95.6  38.2  91.7 | 4.5  38.1  2.7  41.8  8.3 | 0.9  15.9  1.8  16.4  0 | 0  0  0  0.9  0 | 0  0  0  0.9  0 |
| Any solicited systemic AE | 1 | A  B  C  D  E | 0/0  5/50  5/50  25/50  25/50 | 113  116  117  113  117 | 68.1  62.1  69.2  70.8  65.8 | 20.4  22.4  16.2  21.2  23.1 | 10.6  12.1  12.8  6.2  10.3 | 0  3.4  1.7  1.8  0 | 0.9  0  0  0  0.9 |
|  | 2 | A  B  C  D  E | 0/0  5/50  0/0  25/50  0/0 | 109  113  113  110  108 | 78.0  61.1  77.9  49.1  79.6 | 14.7  19.5  13.3  20.9  13.0 | 6.4  16.8  7.1  25.5  7.4 | 0.9  2.7  1.8  3.6  0 | 0  0  0  0.9  0 |
| Joint pain/ arthralgia | 1 | A  B  C  D  E | 0/0  5/50  5/50  25/50  25/50 | 113  116  117  113  117 | 97.3  94.8  88.9  96.5  91.5 | 2.7  0.9  6.0  3.5  4.3 | 0  3.4  5.1  0  4.3 | 0  0.9  0  0  0 | 0  0  0  0  0 |
|  | 2 | A  B  C  D  E | 0/0  5/50  0/0  25/50  0/0 | 109  113  113  110  108 | 98.2  89.4  96.5  87.3  96.3 | 0  5.3  0.9  4.5  1.9 | 1.8  3.5  2.7  8.2  1.9 | 0  1.8  0  0  0 | 0  0  0  0  0 |
| Fatigue | 1 | A  B  C  D  E | 0/0  5/50  5/50  25/50  25/50 | 113  116  117  113  117 | 82.3  81.0  82.1  89.4  86.3 | 9.7  7.8  8.5  4.4  6.8 | 8.0  9.5  7.7  5.3  6.8 | 0  1.7  1.7  0.9  0 | 0  0  0  0  0 |
|  | 2 | A  B  C  D  E | 0/0  5/50  0/0  25/50  0/0 | 109  113  113  110  108 | 89.9  77.0  88.5  67.3  89.8 | 5.5  8.0  6.2  10.0  3.7 | 3.7  15.0  4.4  20.0  6.5 | 0.9  0  0.9  2.7  0 | 0  0  0  0  0 |
| Malaise | 1 | A  B  C  D  E | 0/0  5/50  5/50  25/50  25/50 | 113  116  117  113  117 | 92.0  84.5  88.9  95.6  89.7 | 2.7  8.6  6.0  2.7  7.7 | 4.4  5.2  3.4  0.9  2.6 | 0  1.7  0  0  0 | 0.9  0  0  0  0 |
|  | 2 | A  B  C  D  E | 0/0  5/50  0/0  25/50  0/0 | 109  113  113  110  108 | 92.7  79.6  93.8  83.6  96.3 | 4.6  7.1  2.7  2.7  2.8 | 2.8  12.4  2.7  11.8  0 | 0  0  0  0  0 | 0  0  0  0  0 |
| Temperature/ Fever | 1 | A  B  C  D  E | 0/0  5/50  5/50  25/50  25/50 | 112  116  117  113  117 | 97.3  96.6  97.4  98.2  98.3 | 0.9  1.7  1.7  0  0 | 1.8  0  0.9  0.9  0.9 | 0  1.7  0  0.9  0 | 0  0  0  0  0.9 |
|  | 2 | A  B  C  D  E | 0/0  5/50  0/0  25/50  0/0 | 109  112  113  108  107 | 99.1  98.2  100.0  96.3  99.1 | 0.9  1.8  0  2.8  0 | 0  0  0  0.9  0.9 | 0  0  0  0  0 | 0  0  0  0  0 |
| Headache | 1 | A  B  C  D  E | 0/0  5/50  5/50  25/50  25/50 | 113  116  117  113  117 | 85.0  83.6  89.7  88.5  89.7 | 13.3  12.9  7.7  10.6  9.4 | 1.8  3.4  2.6  0.9  0.9 | 0  0  0  0  0 | 0  0  0  0  0 |
|  | 2 | A  B  C  D  E | 0/0  5/50  0/0  25/50  0/0 | 109  113  113  110  108 | 89.0  95.0  89.4  74.5  88.9 | 10.1  8.8  8.0  12.7  10.2 | 0.9  5.3  1.8  11.8  0.9 | 0  0.9  0.9  0.9  0 | 0  0  0  0  0 |
| Muscle pain/ Myalgia | 1 | A  B  C  D  E | 0/0  5/50  5/50  25/50  25/50 | 113  116  117  113  117 | 92.0  86.2  83.8  89.4  82.1 | 5.3  10.3  13.7  9.7  15.4 | 2.7  2.6  2.6  0.9  2.6 | 0  0.9  0  0  0s | 0  0  0  0  0 |
|  | 2 | A  B  C  D  E | 0/0  5/50  0/0  25/50  0/0 | 109  113  113  110  108 | 99.1  80.5  96.5  70.9  97.2 | 0  8.8  1.8  13.6  0.9 | 0.9  9.7  1.8  14.5  1.9 | 0  0.9  0  0  0 | 0  0  0  0.9  0 |
| Nausea or vomiting | 1 | A  B  C  D  E | 0/0  5/50  5/50  25/50  25/50 | 113  116  117  113  117 | 98.2  94.8  96.6  97.3  97.4 | 1.8  2.6  2.6  2.7  2.6 | 0  2.6  0.9  0  0 | 0  0  0  0  0 | 0  0  0  0  0 |
|  | 2 | A  B  C  D  E | 0/0  5/50  0/0  25/50  0/0 | 109  113  113  110  108 | 99.1  93.8  99.1  92.7  99.1 | 0.9  5.3  0  5.5  0.9 | 0  0.9  0.9  1.8  0 | 0  0  0  0  0 | 0  0  0  0  0 |

*	AE denotes adverse event, , N denotes number of participants assessed.

†	Group A (placebo): 0 µg NVX-CoV2373/0 µg Matrix-M1 on first vaccination; 0 µg NVX-CoV2373/0 µg Matrix-M1 on second vaccination. Group B: 5 µg NVX-CoV2373/50 µg Matrix-M1 on first vaccination; 5 µg NVX-CoV2373/50 µg Matrix-M1 on second vaccination. Group C: 5 µg NVX-CoV2373/50 µg Matrix-M1 on first vaccination; 0 µg NVX-CoV2373/0 µg Matrix-M1 on second vaccination. Group D: 25 µg NVX-CoV2373/50 µg Matrix-M1 on first vaccination; 25 µg NVX-CoV2373/50 µg Matrix-M1 on second vaccination. Group E: 25 µg NVX-CoV2373/50 µg Matrix-M1 on first vaccination; 0 µg NVX-CoV2373/0 µg Matrix-M1 on second vaccination.

##### Table S5. Treatment-Emergent Adverse Events by System Organ Class and Preferred Term Reported in 1% or More Participants in the Total Group Through 35 Days After First Vaccination (Safety Analysis Set).*

| **Group**  AGEU | **A** | **B** | **C** | **D** | **E** | **Total**  **1283** |
| --- | --- | --- | --- | --- | --- | --- |
| **NVX-CoV2373 Dose 1/2 (µg)** | **0/0** | **5/5** | **5/0** | **25/25** | **25/0** |  |
| **Matrix-M1 Dose 1/2 (µg)** | **0/0** | **50/50** | **50/0** | **50/50** | **50/0** |  |
| **System Organ Class/Preferred Term† N** | **255** | **258** | **256** | **259** | **255** |  |
| Any TEAE‡ | 42 (16.5) | 51 (19.8) | 35 (13.7) | 52 (20.1) | 43 (16.9) | 223 (17.4) |
| Infections and infestations | 9 (3.5) | 8 (3.1) | 7 (2.7) | 10 (3.9) | 6 (2.4) | 40 (3.1) |
| Urinary tract infection | 2 (0.8) | 2 (0.8) | 1 (0.4) | 2 (0.8) | 3 (1.2) | 10 (0.8) |
| Injury, poisoning and procedural complications | 7 (2.7) | 7 (2.7) | 7 (2.7) | 5 (1.9) | 6 (2.4) | 32 (2.5) |
| Gastrointestinal disorders | 7 (2.7) | 9 (3.5) | 3 (1.2) | 6 (2.3) | 6 (2.4) | 31 (2.4) |
| General disorders and administration site conditions | 6 (2.4) | 7 (2.7) | 1 (0.4) | 13 (5.0) | 2 (0.8) | 29 (2.3) |
| Injection site pruritus | 0 | 3 (1.2) | 0 | 5 (1.9) | 0 | 8 (0.6) |
| Musculoskeletal and connective tissue disorders | 5 (2.0) | 8 (3.1) | 5 (2.0) | 5 (1.9) | 4 (1.6) | 27 (2.1) |
| Arthralgia | 2 (0.8) | 3 (1.2) | 0 | 1 (0.4) | 1 (0.4) | 7 (0.5) |
| Nervous system disorders | 4 (1.6) | 5 (1.9) | 4 (1.6) | 6 (2.3) | 2 (0.8) | 21 (1.6) |
| Headache | 2 (0.8) | 3 (1.2) | 1 (0.4) | 2 (0.8) | 1 (0.4) | 9 (0.7) |
| Respiratory, thoracic and mediastinal disorders | 3 (1.2) | 4 (1.6) | 3 (1.2) | 5 (1.9) | 4 (1.6) | 19 (1.5) |
| Skin and subcutaneous tissue disorders | 2 (0.8) | 3 (1.2) | 2 (0.8) | 3 (1.2) | 8 (3.1) | 18 (1.4) |
| Blood and lymphatic system  disorders | 1 (0.4) | 3 (1.2) | 1 (0.4) | 2 (0.8) | 2 (0.8) | 9 (0.7) |
| Lymphadenopathy | 1 (0.4) | 3 (1.2) | 1 (0.4) | 1 (0.4) | 2 (0.8) | 8 (0.6) |
| Vascular disorders | 2 (0.8) | 2 (0.8) | 1 (0.4) | 0 | 3 (1.2) | 8 (0.6) |
| Hypertension | 1 (0.4) | 2 (0.8) | 0 | 0 | 3 (1.2) | 6 (0.5) |

* TEAE denotes treatment-emergent adverse event.

† Adverse events were coded using MedDRA Version 23.0.

‡ Based on all TEAEs.

##### Table S6. Primary Endpoint Analysis ‒ Geometric Mean Titer IgG Responses (ELISA Units) for Anti-S Protein IgG Antibody Levels for SARS-CoV-2 rS Protein Antigen With Matrix M1 Adjuvant in Participants 18 to 84 Years of Age at Day 35 (PP Immunogenicity Analysis Set).

| **Vaccine Group** | **Group A** | **Group B** | **Group C** | **Group D** | **Group E** |
| --- | --- | --- | --- | --- | --- |
| **NVX-CoV2373 Dose 1/2 (µg)** | **0/0** | **5/5** | **5/0** | **25/25** | **25/0** |
| **Matrix-M1 Dose 1/2 (µg)** | **0/0** | **50/50** | **50/0** | **50/50** | **50/0** |
| Day 35 |  |  |  |  |  |
| n1 | 238 | 240 | 241 | 236 | 243 |
| GMT (EU/mL) | 126.1 | 44420.9 | 894.0 | 46459.3 | 1951.3 |
| 95% CI | 114.0, 139.4 | 37929.1, 52023.8 | 744.1, 1074.0 | 40839.4, 52852.5 | 1658.3, 2296.1 |
| GMFR referencing Day 0 | 1.0 | 385.6 | 7.4 | 384.9 | 15.4 |
| 95% CI | 1.0, 1.1 | 325.5, 456.8 | 6.3, 8.7 | 334.7, 442.7 | 13.3, 17.9 |
| SCR ≥ 4-fold increase, n2/n1 (%) | 3/238 (1.3) | 236/240 (98.3) | 163/241 (67.6) | 235/236 (99.6) | 211/243 (86.8) |
| 95% CI | 0.3, 3.6 | 95.8, 99.5 | 61.3, 73.5 | 97.7, 100.0 | 81.9, 90.8 |

Abbreviations: CI = confidence interval; COVID-19 = coronavirus disease 2019; ELISA = enzyme-linked immunosorbent assay; EU = ELISA units; GMFR = geometric mean fold rise; GMT = geometric mean titer; IgG = immunoglobulin G; LLOQ = lower limit of quantification; n1 = number of participants in the PP‑Immunogenicity Set within each visit with non-missing data; n2 = the number of participants who reported the event; n/a = not applicable; PP = per-protocol; SARS-CoV-2 rS = severe acute respiratory syndrome coronavirus 2 recombinant spike protein nanoparticle vaccine; SCR = seroconversion rate.

Note: LLOQ = 200 EU/mL. Percentages were calculated as (n2/n1) × 100. Titer values less than LLOQ were replaced by 0.5 × LLOQ. The 95th percentile was calculated relative to placebo participants who remain COVID-19 free at the applicable visit. The 95% CI for GMT and GMFR were calculated based on the t-distribution of the log-transformed values, then back transformed to the original scale for presentation. The 95% CI for SCR was calculated using the exact Clopper-Pearson method. 95% CI for the difference of SCR was calculated using the method of Miettinen and Nurminen.

##### Table S7. Geometric Mean Titer IgG Responses (ELISA Units) for Anti-S Protein IgG Antibody Levels for SARS-CoV-2 rS Protein Antigen With Matrix M1 Adjuvant in Participants 18 to 59 Years of Age at Baseline, Day 21 and Day 35 (PP Analysis Set).

| **Vaccine Group** | **Group A** | **Group B** | **Group C** | **Group D** | **Group E** |
| --- | --- | --- | --- | --- | --- |
| **NVX-CoV2373 Dose 1/2 (µg)** | **0/0** | **5/5** | **5/0** | **25/25** | **25/0** |
| **Matrix-M1 Dose 1/2 (µg)** | **0/0** | **50/50** | **50/0** | **50/50** | **50/0** |
| Day 0 |  |  |  |  |  |
| n1 | 138 | 137 | 140 | 143 | 139 |
| GMT (EU/mL) | 116.5 | 119.1 | 123.4 | 125.2 | 119.6 |
| 95% CI | (105.2,128.9) | (105.6,134.2) | (111.3,136.8) | (110.7,141.7) | (107.6, 132.9) |
| Day 21 |  |  |  |  |  |
| n1 | 137 | 135 | 139 | 142 | 133 |
| GMT (EU/mL) | 121.0 | 1374.0 | 1457.8 | 3155.7 | 2897.6 |
| 95% CI | (108.7, 134.7) | (1109.4, 1701.7) | (1170.6,  1815.4) | (2543.3,  3915.7) | (2341.4,  3586.0) |
| GMFR referencing Day 0 | 1.0 | 11.5 | 11.8 | 25.2 | 24.0 |
| 95% CI | (1.0, 1.1) | (9.4, 14.1) | (9.6, 14.5) | (20.5, 30.9) | (19.7, 29.3) |
| SCR ≥ 4-fold increase, n2/n1 (%) | 2/137 (1.5) | 106/135 (78.5) | 110/139 (79.1) | 133/142 (93.7) | 123/133 (92.5) |
| 95% CI | (0.2, 5.2) | (70.6, 85.1) | (71.4, 85.6) | (88.3, 97.1) | (86.6, 96.3) |
| Day 35 |  |  |  |  |  |
| n1 | 135 | 127 | 134 | 137 | 133 |
| GMT (EU/mL) | 123.9 | 65019.1 | 1493.4 | 58773.8 | 2644.3 |
| 95% CI | 109.0, 140.9 | 55484.8, 76191.9 | 1206.1, 1849.2 | 51611.7, 66929.8 | 2175.6, 3213.9 |
| GMFR referencing Day 0 | 1.1 | 538.6 | 12.1 | 464.7 | 21.9 |
| 95% CI | 1.0, 1.2 | 442.1, 656.2 | 9.9, 14.9 | 395.2, 546.4 | 18.3, 26.3 |
| SCR ≥ 4-fold increase, n2/n1 (%) | 2/135 (1.5) | 126/127 (99.2) | 109/134 (81.3) | 137/137 (100.0) | 125/133 (94.0) |
| 95% CI | 0.2, 5.2 | 95.7, 100.0 | 73.7, 87.5 | 97.3, 100.0 | 88.5, 97.4 |

Abbreviations: CI = confidence interval; COVID-19 = coronavirus disease 2019; ELISA = enzyme-linked immunosorbent assay; EU = ELISA units; GMFR = geometric mean fold rise; GMT = geometric mean titer; IgG = immunoglobulin G; LLOQ = lower limit of quantification; n1 = number of participants in the PP‑Immunogenicity Set within each visit with non-missing data; n2 = the number of participants who reported the event; n/a = not applicable; PP = per-protocol; SARS-CoV-2 rS = severe acute respiratory syndrome coronavirus 2 recombinant spike protein nanoparticle vaccine; SCR = seroconversion rate.

Note: LLOQ = 200 EU/mL. Percentages were calculated as (n2/n1) × 100. Titer values less than LLOQ were replaced by 0.5 × LLOQ. The 95th percentile was calculated relative to placebo participants who remain COVID-19 free at the applicable visit. The 95% CI for GMT and GMFR were calculated based on the t-distribution of the log-transformed values, then back transformed to the original scale for presentation. The 95% CI for SCR was calculated using the exact Clopper-Pearson method. 95% CI for the difference of SCR was calculated using the method of Miettinen and Nurminen.

##### Table S8. Microneutralization50 Titers Specific for SARS-CoV-2 Wild-Type for SARS-CoV-2 rS Protein Antigen With Matrix M1 Adjuvant in Participants 18 to 84 Years of Age at Day 35 (PP Analysis Set).

| **Vaccine Group** | **Group A** | **Group B** | **Group C** | **Group D** | **Group E** |
| --- | --- | --- | --- | --- | --- |
| **NVX-CoV2373 Dose 1/2 (µg)** | **0/0** | **5/5** | **5/0** | **25/25** | **25/0** |
| **Matrix-M1 Dose 1/2 (µg)** | **0/0** | **50/50** | **50/0** | **50/50** | **50/0** |
| Day 0 |  |  |  |  |  |
| n1 | 51 | 51 | 57 | 49 | 48 |
| GMT (MN50) | 10.0 | 10.6 | 10.0 | 10.0 | 10.0 |
| 95% CI | (10.0, 10.0) | (9.5, 11.8) | (10.0, 10.0) | (10.0, 10.0) | (10.0, 10.0) |
| Day 21 |  |  |  |  |  |
| n1 | 21 | 21 | 22 | 21 | 20 |
| GMT (MN50) | 10.0 | 40.0 | 30.1 | 67.8 | 44.4 |
| 95% CI | (10.0, 10.0) | (20.3, 78.7) | (20.2, 44.9) | (37.3, 123.3) | (22.9, 85.8) |
| GMFR referencing Day 0 | 1.0 | 3.5 | 3.0 | 6.8 | 4.4 |
| 95% CI | (1.0, 1.0) | (2.0, 6.0) | (2.0, 4.5) | (3.7, 12.3) | (2.3, 8.6) |
| SCR ≥ 4-fold increase, n2/n1 (%) | 0/21 (0.0) | 9/51 (42.9) | 11/22 (50.0) | 15/21 (71.4) | 12/20 (60.0) |
| 95% CI | (0.0, 16.1) | (21.8, 66.0) | (28.2, 71.8) | (47.8, 88.7) | (36.1, 80.9) |
| Day 35 |  |  |  |  |  |
| n1 | 51 | 49 | 55 | 49 | 47 |
| GMT (MN50) | 10.7 | 1433 | 20.8 | 1335.5 | 37.7 |
| 95% CI | (9.3, 12.3) | (978.2, 2099.4) | (16.3, 26.5) | ( 973.8, 1831.6) | (26.9, 52.8) |
| GMFR referencing Day 0 | 1.1 | 143.3 | 2.1 | 133.5 | 3.8 |
| 95% CI | (0.9, 1.2) | (97.8, 209.9) | (1.6, 2.6) | (97.4, 183.2) | (2.7, 5.3) |
| SCR ≥ 4-fold increase, n2/n1 (%) | 1/51 (2.0) | 49/49 (100.0) | 19/55 (34.5) | 48/49 (98.0) | 26/47 (55.3) |
| 95% CI | (0.0, 10.4) | (92.7, 100.0) | (22.2, 48.6) | (89.1, 99.9) | (40.1, 69.8) |

Abbreviations: CI = confidence interval; COVID-19 = coronavirus disease 2019; MN50 = microneutralization titer expressed as the reciprocal of the highest dilution at which 50% of the replicate wells are protected from infection; GMFR = geometric mean fold rise; GMT = geometric mean titer; LLOQ = lower limit of quantification; n1 = number of participants in the PP Analysis Set within each visit with non-missing data; n2 = the number of participants who reported the event; n/a = not applicable; PP = per-protocol; SARS-CoV-2 = severe acute respiratory syndrome coronavirus 2; SARS-CoV-2 rS = severe acute respiratory syndrome coronavirus 2 recombinant spike nanoparticle vaccine; SCR = seroconversion rate.

Note: MN50 titer LLOQ = 20. Percentages were calculated as (n2/n1) × 100. Titer values less than LLOQ were replaced by 0.5 × LLOQ. The 95th percentile was calculated relative to placebo participants who remain COVID-19 free at the applicable visit. The 95% CI for GMT and GMFR were calculated based on the t-distribution of the log-transformed values, then back transformed to the original scale for presentation. The 95% CI for SCR was calculated using the exact Clopper-Pearson method. 95% CI for the difference of SCR was calculated using the method of Miettinen and Nurminen.

##### Table S9. Geometric Mean Titer IgG Responses (ELISA Units) for Anti-S Protein IgG Antibody Levels for SARS-CoV-2 rS Protein Antigen With Matrix M1 Adjuvant in Participants 60 to 84 Years of Age at Baseline, Day 21 and Day 35 (PP Analysis Set).

| **Vaccine Group** | **Group A** | **Group B** | **Group C** | **Group D** | **Group E** |
| --- | --- | --- | --- | --- | --- |
| **NVX-CoV2373 Dose 1/2 (µg)** | **0/0** | **5/5** | **5/0** | **25/25** | **25/0** |
| **Matrix-M1 Dose 1/2 (µg)** | **0/0** | **50/50** | **50/0** | **50/50** | **50/0** |
| Day 0 |  |  |  |  |  |
| n1 | 111 | 117 | 117 | 112 | 117 |
| GMT (EU/mL) | 125.9 | 113.0 | 117.6 | 115.5 | 134.0 |
| 95% CI | (109.6, 144.6) | (102.9, 124.0) | (103.6, 133.5) | (106.0, 125.8) | (116.7, 153.9) |
| Day 21 |  |  |  |  |  |
| n1 | 108 | 116 | 114 | 107 | 114 |
| GMT (EU/mL) | 120.0 | 456.4 | 403.6 | 746.3 | 1214.3 |
| 95% CI | (106.0, 135.9) | (361.9, 575.6) | (308.8, 527.4) | (592.2, 940.5) | (932.7, 1580.8) |
| GMFR referencing Day 0 | 1.0 | 4.0 | 3.5 | 6.5 | 9.0 |
| 95% CI | (0.9, 1.0) | (3.3, 5.0) | (2.8, 4.3) | (5.2, 8.1) | (7.1, 11.4) |
| SCR ≥ 4-fold increase, n2/n1 (%) | 0/108 (0.0) | 53/116 (45.7) | 46/114 (40.4) | 73/107 (68.2) | 92/114 (80.7) |
| 95% CI | (0.0, 3.4) | (36.4, 55.2) | (31.3, 49.9) | (58.5, 76.9) | (72.3, 87.5) |
| Day 35 |  |  |  |  |  |
| n1 | 107 | 114 | 112 | 105 | 110 |
| GMT (EU/mL) | 127.8 | 28136.6 | 457.9 | 32871.2 | 1347.5 |
| 95% CI | 109.2, 149.6 | 21616.6, 36623.3 | 351.1, 597.1 | 26189.5, 41257.5 | 1041.5, 1743.5 |
| GMFR referencing Day 0 | 1.0 | 257.7 | 3.9 | 286.3 | 9.9 |
| 95% CI | 0.9, 1.1 | 197.1, 336.9 | 3.1, 4.9 | 226.6, 361.8 | 7.9, 12.3 |
| SCR ≥ 4-fold increase, n2/n1 (%) | 1/107 (0.9) | 111/114 (97.4) | 55/112 (49.1) | 104/105 (99.0) | 86/110 (78.2) |
| 95% CI | 0.0, 5.1 | 92.5, 99.5 | 39.5, 58.7 | 94.8, 100.0 | 69.3, 85.5 |

Abbreviations: CI = confidence interval; COVID-19 = coronavirus disease 2019; ELISA = enzyme-linked immunosorbent assay; EU = ELISA units; GMFR = geometric mean fold rise; GMT = geometric mean titer; IgG = immunoglobulin G; LLOQ = lower limit of quantification; n1 = number of participants in the PP‑Immunogenicity Set within each visit with non-missing data; n2 = the number of participants who reported the event; n/a = not applicable; PP = per-protocol; SARS-CoV-2 rS = severe acute respiratory syndrome coronavirus 2 recombinant spike protein nanoparticle vaccine; SCR = seroconversion rate.

Note: LLOQ = 200 EU/mL. Percentages were calculated as (n2/n1) × 100. Titer values less than LLOQ were replaced by 0.5 × LLOQ. The 95th percentile was calculated relative to placebo participants who remain COVID-19 free at the applicable visit. The 95% CI for GMT and GMFR were calculated based on the t-distribution of the log-transformed values, then back transformed to the original scale for presentation. The 95% CI for SCR was calculated using the exact Clopper-Pearson method. 95% CI for the difference of SCR was calculated using the method of Miettinen and Nurminen.

##### Table S10. Microneutralization50 Titers Specific for SARS-CoV-2 Wild-Type for SARS-CoV-2 rS Protein Antigen With Matrix M1 Adjuvant in Participants 18 to 59 Years of Age at Day 35 (PP Analysis Set).

| **Vaccine Group** | **Group A** | **Group B** | **Group C** | **Group D** | **Group E** |
| --- | --- | --- | --- | --- | --- |
| **NVX-CoV2373 Dose 1/2 (µg)** | **0/0** | **5/5** | **5/0** | **25/25** | **25/0** |
| **Matrix-M1 Dose 1/2 (µg)** | **0/0** | **50/50** | **50/0** | **50/50** | **50/0** |
| Day 0 |  |  |  |  |  |
| n1 | 26 | 24 | 31 | 23 | 24 |
| GMT (MN50) | 10.0 | 10.0 | 10.0 | 10.0 | 10.0 |
| 95% CI | (10.0, 10.0) | (10.0, 10.0) | (10.0, 10.0) | (10.0, 10.0) | (10.0, 10.0) |
| Day 21 |  |  |  |  |  |
| n1 | 11 | 8 | 12 | 11 | 10 |
| GMT (MN50) | 10.0 | 36.7 | 44.9 | 132.4 | 60.6 |
| 95% CI | (10.0, 10.0) | (12.3, 109.4) | (26.5, 75.9) | (64.2, 273.2) | (17.9, 205.2) |
| GMFR referencing Day 0 | 1.0 | 3.7 | 4.5 | 13.2 | 6.1 |
| 95% CI | (1.0, 1.0) | (1.2, 10.9) | (2.7, 7.6) | (6.4, 27.3) | (1.8, 20.5) |
| SCR ≥ 4-fold increase, n2/n1 (%) | 0/11 (0.0) | 3/8 (37.5) | 8/12 (66.7) | 10/11 (90.9) | 7/10 (70.0) |
| 95% CI | (0.0, 28.5) | (8.5, 75.5) | (34.9, 90.1) | (58.7, 99.8) | ( 34.8, 93.3) |
| Day 35 |  |  |  |  |  |
| n1 | 26 | 23 | 29 | 23 | 23 |
| GMT (MN50) | 11.4 | 2200.8 | 26.6 | 1783.1 | 52.5 |
| 95% CI | (8.7, 15.0) | (1342.6, 3607.5) | (18.1, 39.3) | (1191.9, 2667.7) | (31.3, 88.0) |
| GMFR referencing Day 0 | 1.1 | 220.1 | 2.7 | 178.3 | 5.2 |
| 95% CI | (0.9, 1.5) | (134.3, 360.7) | (1.8, 3.9) | (119.2, 266.8) | (3.1, 8.8) |
| SCR ≥ 4-fold increase, n2/n1 (%) | 1/26 (3.8) | 23/23 (100.0) | 13/29 (44.8) | 23/23 (100.0) | 15/23 (65.2) |
| 95% CI | (0.1, 19.6) | (85.2, 100.0) | (26.4, 64.3) | (85.2, 100.0) | (42.7, 83.6) |

Abbreviations: CI = confidence interval; COVID-19 = coronavirus disease 2019; MN50 = microneutralization titer expressed as the reciprocal of the highest dilution at which 50% of the replicate wells are protected from infection; GMFR = geometric mean fold rise; GMT = geometric mean titer; LLOQ = lower limit of quantification; n1 = number of participants in the PP Analysis Set within each visit with non-missing data; n2 = the number of participants who reported the event; n/a = not applicable; PP = per-protocol; SARS-CoV-2 = severe acute respiratory syndrome coronavirus 2; SARS-CoV-2 rS = severe acute respiratory syndrome coronavirus 2 recombinant spike nanoparticle vaccine; SCR = seroconversion rate.

Note: MN50 titer LLOQ = 20. Percentages were calculated as (n2/n1) × 100. Titer values less than LLOQ were replaced by 0.5 × LLOQ. The 95th percentile was calculated relative to placebo participants who remain COVID-19 free at the applicable visit. The 95% CI for GMT and GMFR were calculated based on the t-distribution of the log-transformed values, then back transformed to the original scale for presentation. The 95% CI for SCR was calculated using the exact Clopper-Pearson method. 95% CI for the difference of SCR was calculated using the method of Miettinen and Nurminen.

##### Table S11. Microneutralization50 Titers Specific for SARS-CoV-2 Wild-Type for SARS-CoV-2 rS Protein Antigen With Matrix M1 Adjuvant in Participants 60-84 Years of Age at Day 35 (PP Analysis Set).

| **Vaccine Group** | **Group A** | **Group B** | **Group C** | **Group D** | **Group E** |
| --- | --- | --- | --- | --- | --- |
| **NVX-CoV2373 Dose 1/2 (µg)** | **0/0** | **5/5** | **5/0** | **25/25** | **25/0** |
| **Matrix-M1 Dose 1/2 (µg)** | **0/0** | **50/50** | **50/0** | **50/50** | **50/0** |
| Day 0 |  |  |  |  |  |
| n1 | 25 | 27 | 26 | 26 | 24 |
| GMT (MN50) | 10.0 | 11.1 | 10.0 | 10.0 | 10.0 |
| 95% CI | (10.0, 10.0) | (9.0, 13.7) | (10.0, 10.0) | (10.0, 10.0) | (10.0, 10.0) |
| Day 21 |  |  |  |  |  |
| n1 | 10 | 13 | 10 | 10 | 10 |
| GMT (MN50) | 10.0 | 42.2 | 18.7 | 32.5 | 32.5 |
| 95% CI | (10.0, 10.0) | (15.7,  113.5) | (10.8,  32.2) | (14.0,  75.6) | (15.5,  68.2) |
| GMFR referencing Day 0 | 1.0 | 3.4 | 1.9 | 3.2 | 3.2 |
| 95% CI | (1.0, 1.0) | (1.7, 6.9) | (1.1, 3.2) | (1.4, 7.6) | (1.5, 6.8) |
| SCR ≥ 4-fold increase, n2/n1 (%) | 0/10 (0.0) | 6/13 (46.2) | 3/10 (30.0) | 5/10 (50.0) | 5/10 (50.0) |
| 95% CI | (0.0, 30.8) | (19.2, 74.9) | (6.7, 65.2) | (18.7, 81.3) | (18.7, 81.3) |
| Day 35 |  |  |  |  |  |
| n1 | 25 | 26 | 26 | 26 | 24 |
| GMT (MN50) | 10.0 | 980.5 | 15.7 | 1034.2 | 27.5 |
| 95% CI | (10.0, 10.0) | (559.8, 1717.1) | (12.1, 20.4) | (639.8, 1671.6) | (17.9, 42.3) |
| GMFR referencing Day 0 | 1.0 | 98.0 | 1.6 | 103.4 | 2.7 |
| 95% CI | (1.0, 1.0) | (56.0, 171.7) | (1.2, 2.0) | (64.0, 167.2) | (1.8, 4.2) |
| SCR ≥ 4-fold increase, n2/n1 (%) | 0/25 (0.0) | 26/26 (100.0) | 6/26 (23.1) | 25/26 (96.2) | 11/24 (45.8) |
| 95% CI | (0.0, 13.7) | (86.8, 100.0) | (9.0, 43.6) | (80.4, 99.9) | (25.6, 67.2) |

Abbreviations: CI = confidence interval; COVID-19 = coronavirus disease 2019; MN50 = microneutralization titer expressed as the reciprocal of the highest dilution at which 50% of the replicate wells are protected from infection; GMFR = geometric mean fold rise; GMT = geometric mean titer; LLOQ = lower limit of quantification; n1 = number of participants in the PP Analysis Set within each visit with non-missing data; n2 = the number of participants who reported the event; n/a = not applicable; PP = per-protocol; SARS-CoV-2 = severe acute respiratory syndrome coronavirus 2; SARS-CoV-2 rS = severe acute respiratory syndrome coronavirus 2 recombinant spike nanoparticle vaccine; SCR = seroconversion rate.

Note: MN50 titer LLOQ = 20. Percentages were calculated as (n2/n1) × 100. Titer values less than LLOQ were replaced by 0.5 × LLOQ. The 95th percentile was calculated relative to placebo participants who remain COVID-19 free at the applicable visit. The 95% CI for GMT and GMFR were calculated based on the t-distribution of the log-transformed values, then back transformed to the original scale for presentation. The 95% CI for SCR was calculated using the exact Clopper-Pearson method. 95% CI for the difference of SCR was calculated using the method of Miettinen and Nurminen.
